# Phenomic environment-wide association study (PheEWAS) models complexity in the exposome

**DOI:** 10.1101/2025.06.13.25329592

**Authors:** Nicole E. Palmiero, Tomás González Zarzar, Kristin Passero, Jiayan Zhou, Dana C. Crawford, Molly A. Hall

**Affiliations:** Department of Genetics, Perelman School of Medicine, University of Pennsylvania, Philadelphia, PA, USA; Neurology Department, School of Medicine, Pontificia Universidad Católica de Chile, Santiago, Chile; Institute for Biological and Medical Engineering, Pontificia Universidad Católica de Chile, Santiago, Chile; Virginia Institute of Psychiatric and Behavioral Genetics, Virginia Commonwealth University, Richmond, VA, USA; Department of Medicine, Division of Cardiovascular Medicine, Stanford University School of Medicine, Stanford, CA, USA; Departments of Population and Quantitative Health Sciences, and Genetics and Genome Sciences, Cleveland Institute for Computational Biology, Case Western Reserve University (CWRU), Cleveland, OH, USA

**Author notes:** Please contact Molly A. Hall at for requests of correspondence and materials.

## Abstract

Phenome-wide association studies (PheWAS) have successfully identified genomic-based interrelationships between phenotypes but seldom consider environmental exposures. Here, in a phenomic environment-wide association study (PheEWAS), we interrogated relationships between 326 exposures and 55 phenotypes for ∼19,000 participants of the National Health and Nutrition Examination Survey (NHANES). Linear regression models adjusted for age, sex, socioeconomic status, BMI, race/ethnicity, and survey year identified and replicated 106 significant exposure–phenotype associations after Bonferroni correction. The top association was for alpha-tocopherol (vitamin E) with triglycerides (Discovery p = 1.16 × 10⁻¹¹; Replication p = 8.05 × 10⁻¹³). The exposure retinol (vitamin A) had the largest number of individual replicating associations (14 phenotypes including total calcium, iron-binding capacity, ferritin, albumin, transferrin saturation, creatinine, gamma-glutamyl transferase, triglycerides, uric acid, alkaline phosphatase, hemoglobin, and blood urea nitrogen). The phenotype with the greatest number of exposure associations was homocysteine (associated with thiamine; alpha- and gamma-tocopherol; dietary fiber, protein, and potassium; riboflavin; cotinine; folate; phosphorus; cadmium; iron intake; supplement count; and niacin). A race/ethnicity-stratified analysis revealed 11 unique population-specific associations. Our findings demonstrate PheEWAS a method to provide new details on the complexity of the exposome at the level of the phenome

## INTRODUCTION

The association of genomic variants with multiple phenotypes using phenome-wide association studies (PheWAS) has identified pleiotropic associations and other phenomic relationships using genetic data from epidemiological studies and clinical trials^1–4^. Applying a phenome-wide approach to genomics research broadens the context of genetic findings and uncovers new information on the development of human diseases^5^. Although exposome research has typically explored phenotypes individually, phenome-wide exposure studies have provided new information on correlations between multiple phenotypes and a single environmental exposure^6,7^. Here, we present a novel approach for investigating the exposome (i.e., the lifetime environmental exposures of an individual): phenomic environment-wide association studies (PheEWAS).

The potential to detect pleiotropy through cross-phenotype associations is a key advantage of PheWAS, as it identifies genetic variants with wide-reaching health impacts. For example, PheWAS have identified genes simultaneously related to lipoprotein metabolism, vitamin E levels, folate levels, and triglycerides^2^. However, exogenous factors such as pollutants, allergens, toxicants, or infectious agents also contribute to human traits and disease^8,9^. Environment-wide association studies (EWAS), which are used to determine the effect of the exposome on human health, have identified exposures associated with type 2 diabetes, body mass index (BMI), blood pressure, and others^10,11^. Identifying exposures with multiple phenotypic effects (i.e., “cross-phenotype exposures”) can reveal critical environmental risk factors with far-reaching impacts on disease outcomes. For example, exposure to heavy metals contributes to multiple diseases with varied phenotypic effects^12,13^. Extending EWAS to the phenome level offers the opportunity to discover new insights into the far-reaching effects of the environment on human health. PheEWAS tests associations between a wide range of exposures and phenotypes, providing information on the complex network of interconnections between the exposome and phenome.

We developed high-throughput methods to identify associations between the exposome and the phenome, with robust quality control (QC) and defined analysis pipelines using our publicly available software, CLeaning to Analysis: Reproducibility-based Interface for Traits and Exposures (CLARITE)^11^. Using data from the National Health and Nutrition Examination Survey (NHANES)^14^, we evaluated associations between 326 exposures and 55 phenotypes, using data from more than 18,000 participants. The environmental exposures ranged from physical and chemical measurements to survey responses. The phenotypes included biochemical blood sample measurements. Our PheEWAS reveals interrelationships between the exposome and phenome that are important to consider for human health.

## RESULTS

### PheEWAS results

We considered 326 exposures and 55 phenotypes that passed our quality control (QC) pipeline (see Methods) for PheEWAS. There were 1,930 statistically significant associations in the discovery dataset with a false discovery rate (FDR) < 0.1. These were considered for replication, and 686 replicated with the same direction of effect in the replication dataset with FDR < 0.1 (Fig. 1). One hundred and six exposure-phenotype associations demonstrated a Bonferroni-adjusted p-value < 0.05 in both discovery and replication datasets (Table 1). The exposures in these 106 PheEWAS associations were categorized as nutrients (30), food component recall (18), heavy metals (14), smoking behavior (14), smoking in the family (9), smoke exposure measured through cotinine (12), alcohol use (5), pharmaceutical (3), and supplement use (1). The phenotypes were categorized as metabolic products (19), blood proteins (19), white blood cells (WBC) (15), red blood cell (RBC) (14), amino acids (14), enzymes (9), iron measurements (8), and lipids (8). The top exposure-phenotype associations (“d” subscript = discovery and “r” subscript = replication) were alpha-tocopherol (vitamin E) with triglycerides (P_d_ = 6.51 × 10^⁻16^, β_d_ = 0.43; Pᵣ = 4.20 × 10^⁻16^, βᵣ = 0.46), gamma-tocopherol (vitamin E) with triglycerides (P_d_ = 2.46 × 10^⁻14^, β_d_ = 0.37; Pᵣ = 8.54 × 10^⁻16^, βᵣ = 0.37). The 106 Bonferroni-significant associations included 28 unique exposures and 32 unique phenotypes. This shows that many exposures and phenotypes had repeated significant associations. As viewed in the network plot (Fig 2.), the most common significantly associated exposure was retinol, and the most common significantly associated phenotype was homocysteine. The exposure category with the largest number of replicated exposures was nutrients, specifically vitamins (Table 1 & Supplemental Table 1).

**Fig. 1.**
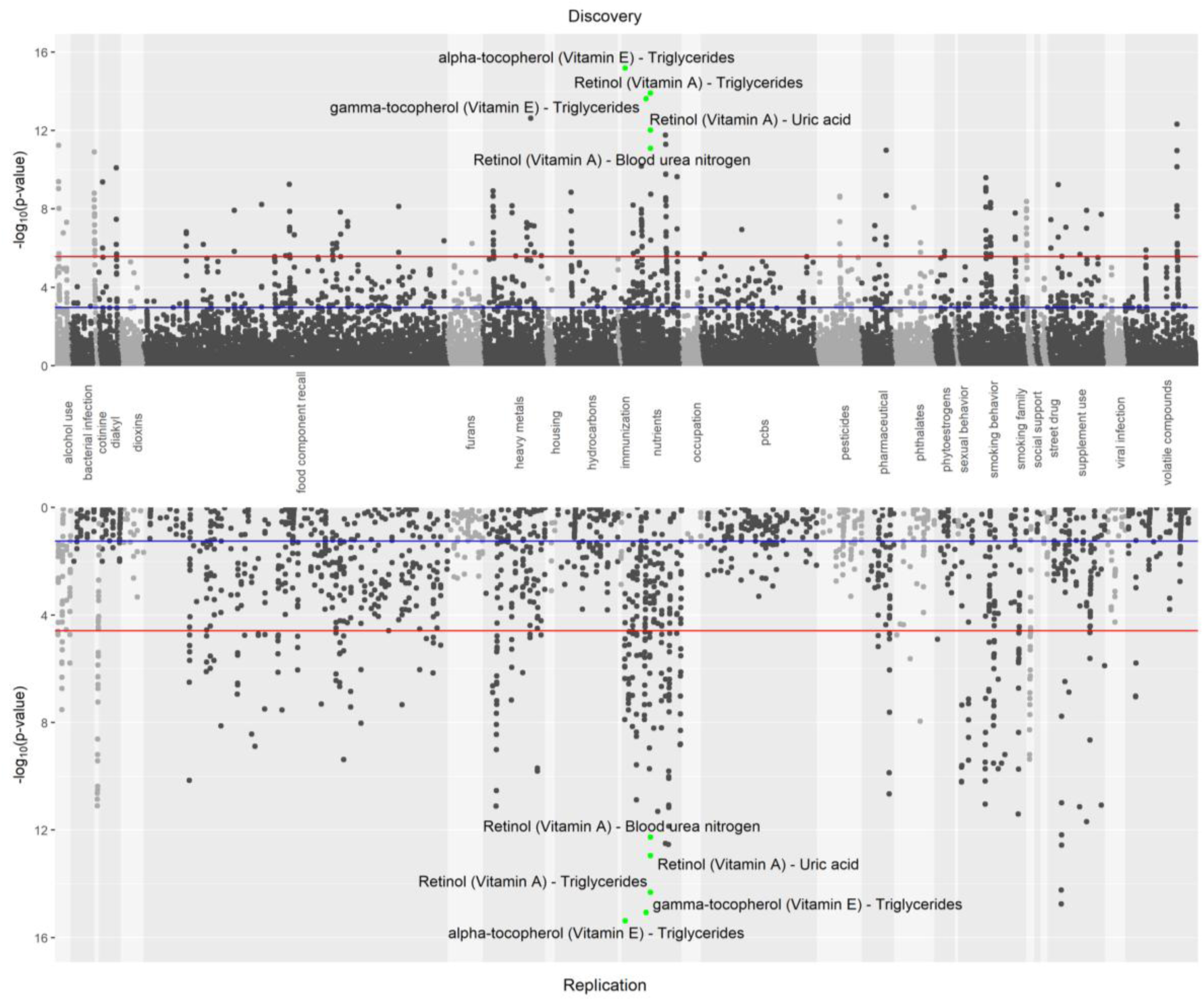
Phenomic environment-wide association study results. Hudson plot of PheEWAS results. The x-axis shows exposure-phenotype associations, ordered by exposure category, and the y-axis shows the −log10(p-value). The red lines denote a Bonferroni-adjusted p-value of 0.05, and the blue lines denote the threshold for a false discovery rate (FDR) of 0.1. The top replicating results in the nutrients category that were significant after FDR adjustment and Bonferroni correction are labeled. The Hudson plot was created using an R package available at https://github.com/anastasia-lucas/hudson67.

**Fig. 2.**
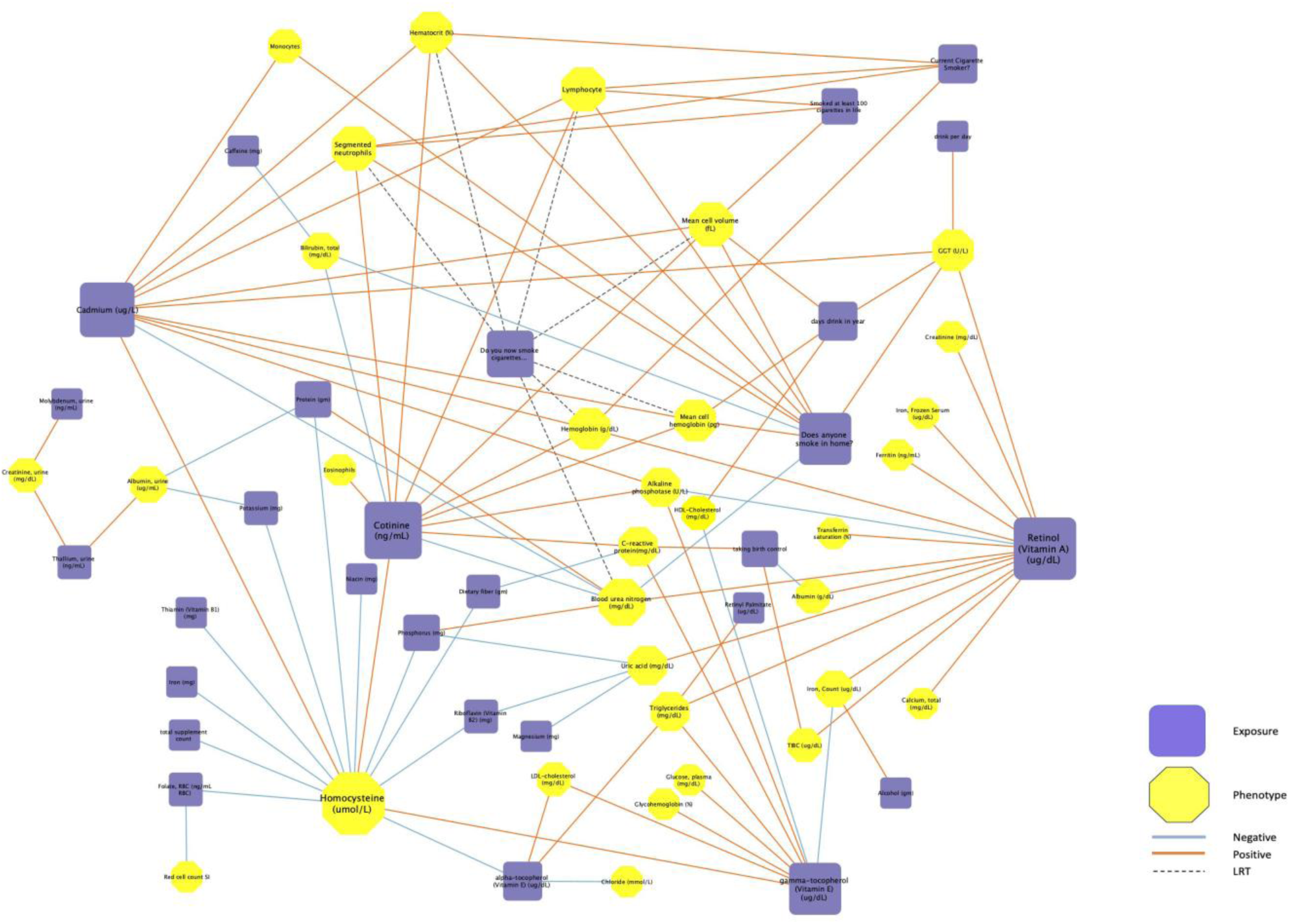
Network-level exposome-phenome associations. A network plot with exposure nodes (purple squares) and phenotype nodes (yellow hexagons) provides a visual description of the various kinds of associations found68. The larger the node the more connections are associated with that node. Line color denotes the type of association. Blue and red connections signify negative and positive GLM coefficients, respectively. The black dashed connections signify a significant LRT because not a single beta value was assigned (See Methods). This plot was generated using Cytoscape software69.

**Table 1:** The top 106 replicating PheEWAS results.

| Exposure | Phenotype | Exposure Category | Phenotype Category | Variable Type | Discovery | Discovery | Replication | Replication |
| --- | --- | --- | --- | --- | --- | --- | --- | --- |
|  |  |  |  |  | Bonferroni p-value | Beta Coefficient | Bonferroni p-value | Beta Coefficient |
| Retinol<br>(vitamin A)<br>(µg/dL) | Albumin (g/dL) | nutrients | biochemistry | continuous | 2.98E-06 | 1.94E-01 | 1.87E-07 | 1.59E-01 |
| Taking birth<br>control | Albumin (g/dL) | pharmaceutical | biochemistry | binary | 1.21E-02 | -4.31E-01 | 4.57E-05 | -5.95E-01 |
| Potassium<br>(mg) | Albumin, urine<br>(µg/mL) | food<br>component<br>recall | biochemistry | continuous | 9.80E-03 | -9.22E-02 | 4.27E-04 | -1.07E-01 |
| Protein (gm) | Albumin, urine<br>(µg/mL) | food<br>component<br>recall | biochemistry | continuous | 4.82E-02 | -8.72E-02 | 3.25E-02 | -7.94E-02 |
| Thallium,<br>urine (ng/mL) | Albumin, urine<br>(µg/mL) | heavy metals | biochemistry | continuous | 2.92E-02 | 2.98E-01 | 3.88E-07 | 3.22E-01 |
| Cadmium<br>(µg/L) | Alkaline c(U/L) | heavy metals | biochemistry | continuous | 1.20E-02 | 9.74E-02 | 1.15E-02 | 6.27E-02 |
| Cotinine<br>(ng/mL) | Alkaline<br>phosphatase<br>(U/L) | cotinine | biochemistry | continuous | 6.10E-05 | 1.02E-01 | 9.57E-04 | 7.97E-02 |
| Gamma-tocopherol<br>(vitamin E)<br>(µg/dL) | Alkaline<br>phosphatase<br>(U/L) | nutrients | biochemistry | continuous | 3.02E-04 | 1.37E-01 | 2.45E-05 | 1.07E-01 |
| Retinol<br>(vitamin A)<br>(µg/dL) | Alkaline<br>phosphatase<br>(U/L) | nutrients | biochemistry | continuous | 4.40E-03 | -8.29E-02 | 1.81E-04 | -9.66E-02 |
| Caffeine (mg) | Bilirubin, total<br>(mg/dL) | food<br>component<br>recall | biochemistry | continuous | 1.19E-02 | -6.39E-02 | 3.17E-02 | -5.98E-02 |
| Cotinine<br>(ng/mL) | Bilirubin, total<br>(mg/dL) | cotinine | biochemistry | continuous | 2.31E-02 | -1.14E-01 | 2.62E-03 | -9.13E-02 |
| Does anyone<br>smoke in<br>home? | Bilirubin, total<br>(mg/dL) | smoking family | biochemistry | binary | 4.30E-02 | -2.26E-01 | 3.80E-05 | -2.36E-01 |
| Cadmium<br>(µg/L) | Blood urea<br>nitrogen (mg/dL) | heavy metals | biochemistry | continuous | 2.60E-02 | -8.99E-02 | 1.04E-03 | -8.73E-02 |
| Cotinine<br>(ng/mL) | Blood urea<br>nitrogen (mg/dL) | cotinine | biochemistry | continuous | 6.12E-05 | -1.22E-01 | 5.99E-08 | -1.21E-01 |
| Do you now<br>smoke<br>cigarettes? | Blood urea<br>nitrogen (mg/dL) | smoking<br>behavior | biochemistry | categorical | 4.90E-03 |  | 1.95E-03 |  |
| Does anyone<br>smoke in<br>home? | Blood urea<br>nitrogen (mg/dL) | smoking family | biochemistry | binary | 1.33E-02 | -2.16E-01 | 1.25E-06 | -2.50E-01 |
| Phosphorus<br>(mg) | Blood urea<br>nitrogen (mg/dL) | food<br>component<br>recall | biochemistry | continuous | 4.07E-02 | 8.54E-02 | 7.08E-04 | 1.04E-01 |
| Protein (gm) | Blood urea<br>nitrogen (mg/dL) | food<br>component<br>recall | biochemistry | continuous | 2.60E-04 | 1.33E-01 | 8.13E-07 | 1.58E-01 |
| Retinol<br>(vitamin A)<br>(µg/dL) | Blood urea<br>nitrogen (mg/dL) | nutrients | biochemistry | continuous | 1.44E-07 | 2.45E-01 | 1.05E-09 | 2.37E-01 |
| Retinol<br>(vitamin A)<br>(µg/dL) | Calcium, total<br>(mg/dL) | nutrients | biochemistry | continuous | 6.71E-05 | 2.24E-01 | 1.34E-08 | 2.01E-01 |
| Alpha-tocopherol<br>(vitamin E)<br>(µg/dL) | Chloride<br>(mmol/L) | nutrients | biochemistry | continuous | 3.37E-02 | -9.52E-02 | 1.85E-04 | -1.31E-01 |
| Cotinine<br>(ng/mL) | C-reactive<br>protein (CRP)<br>(mg/dL) | cotinine | biochemistry | continuous | 1.04E-02 | 9.93E-02 | 5.64E-03 | 8.04E-02 |
| Dietary fiber<br>(gm) | C-reactive<br>protein (CRP)<br>(mg/dL) | food<br>component<br>recall | biochemistry | continuous | 2.69E-02 | -6.82E-02 | 6.28E-03 | -9.90E-02 |
| Gamma-tocopherol<br>(vitamin E)<br>(µg/dL) | C-reactive<br>protein (CRP)<br>(mg/dL) | nutrients | biochemistry | continuous | 7.85E-04 | 9.75E-02 | 4.73E-04 | 1.02E-01 |
| Taking birth<br>control | C-reactive<br>protein (CRP)<br>(mg/dL) | pharmaceutical | biochemistry | binary | 1.86E-07 | 7.94E-01 | 4.20E-08 | 8.24E-01 |
| Retinol<br>(vitamin A)<br>(µg/dL) | Creatinine<br>(mg/dL) | nutrients | biochemistry | continuous | 3.12E-06 | 2.52E-01 | 5.55E-10 | 2.29E-01 |
| Molybdenum,<br>urine (ng/mL) | Creatinine, urine<br>(mg/dL) | heavy metals | biochemistry | continuous | 1.67E-03 | 4.40E-01 | 2.76E-02 | 4.51E-01 |
| Thallium,<br>urine (ng/mL) | Creatinine, urine<br>(mg/dL) | heavy metals | biochemistry | continuous | 1.34E-03 | 6.01E-01 | 2.98E-07 | 6.07E-01 |
| Days drink in<br>year | Direct HDL-cholesterol<br>(mg/dL) | alcohol use | biochemistry | continuous | 1.02E-07 | 2.28E-01 | 5.83E-05 | 1.82E-01 |
| Gamma-tocopherol<br>(vitamin E)<br>(µg/dL) | Direct HDL-cholesterol<br>(mg/dL) | nutrients | biochemistry | continuous | 1.38E-03 | -1.11E-01 | 8.03E-03 | -1.02E-01 |
| Cotinine<br>(ng/mL) | Eosinophils<br>number | cotinine | blood | continuous | 4.42E-02 | 6.58E-02 | 5.02E-04 | 8.35E-02 |
| Retinol<br>(vitamin A)<br>(µg/dL) | Ferritin (ng/mL) | nutrients | biochemistry | continuous | 5.00E-05 | 1.43E-01 | 1.62E-07 | 2.81E-01 |
| Cadmium<br>(µg/L) | GGT (U/L) | heavy metals | biochemistry | continuous | 4.54E-02 | 1.07E-01 | 2.03E-02 | 8.65E-02 |
| Days drink in<br>year | GGT (U/L) | alcohol use | biochemistry | continuous | 1.67E-04 | 1.77E-01 | 3.55E-04 | 1.69E-01 |
| Does anyone<br>smoke in<br>home? | GGT (U/L) | smoking family | biochemistry | binary | 1.66E-03 | 2.71E-01 | 8.38E-04 | 2.10E-01 |
| Drinks per<br>day | GGT (U/L) | alcohol use | biochemistry | continuous | 8.70E-04 | 1.46E-01 | 3.61E-02 | 1.44E-01 |
| Retinol<br>(vitamin A)<br>(µg/dL) | GGT (U/L) | nutrients | biochemistry | continuous | 1.24E-04 | 1.78E-01 | 2.63E-09 | 1.90E-01 |
| Gamma-tocopherol<br>(vitamin E)<br>(µg/dL) | Glucose, plasma<br>(mg/dL) | nutrients | biochemistry | continuous | 1.57E-02 | 1.68E-01 | 5.03E-03 | 1.37E-01 |
| Gamma-tocopherol<br>(vitamin E)<br>(µg/dL) | Glycohemoglobin<br>(%) | nutrients | biochemistry | continuous | 6.13E-04 | 1.78E-01 | 2.43E-03 | 1.29E-01 |
| Cadmium<br>(µg/L) | Hematocrit (%) | heavy metals | blood | continuous | 3.99E-05 | 1.18E-01 | 5.62E-08 | 1.29E-01 |
| Cotinine<br>(ng/mL) | Hematocrit (%) | cotinine | blood | continuous | 4.39E-04 | 1.01E-01 | 4.56E-08 | 1.10E-01 |
| Current<br>cigarette<br>smoker? | Hematocrit (%) | smoking<br>behavior | blood | binary | 1.48E-05 | 2.99E-01 | 3.29E-04 | 2.07E-01 |
| Do you now<br>smoke<br>cigarettes? | Hematocrit (%) | smoking<br>behavior | blood | categorical | 1.00E-04 |  | 1.22E-04 |  |
| Does anyone<br>smoke in<br>home? | Hematocrit (%) | smoking family | blood | binary | 9.98E-03 | 2.11E-01 | 1.51E-04 | 2.28E-01 |
| Cadmium<br>(µg/L) | Hemoglobin<br>(g/dL) | heavy metals | blood | continuous | 2.21E-05 | 1.25E-01 | 1.50E-08 | 1.34E-01 |
| Cotinine<br>(ng/mL) | Hemoglobin<br>(g/dL) | cotinine | blood | continuous | 3.16E-04 | 1.10E-01 | 1.54E-08 | 1.18E-01 |
| Current<br>cigarette<br>smoker? | Hemoglobin<br>(g/dL) | smoking<br>behavior | blood | binary | 2.26E-05 | 3.13E-01 | 3.00E-04 | 2.11E-01 |
| Do you now<br>smoke<br>cigarettes? | Hemoglobin<br>(g/dL) | smoking<br>behavior | blood | categorical | 8.35E-05 |  | 3.29E-05 |  |
| Retinol<br>(vitamin A)<br>(µg/dL) | Hemoglobin<br>(g/dL) | nutrients | blood | continuous | 4.44E-04 | 9.62E-02 | 1.42E-02 | 8.71E-02 |
| Alpha-tocopherol<br>(vitamin E)<br>(µg/dL) | Homocysteine<br>(µmol/L) | nutrients | biochemistry | continuous | 1.86E-03 | -9.80E-02 | 3.09E-06 | -1.14E-01 |
| Cadmium<br>(µg/L) | Homocysteine<br>(µmol/L) | heavy metals | biochemistry | continuous | 4.13E-02 | 8.24E-02 | 5.21E-04 | 9.73E-02 |
| Cotinine<br>(ng/mL) | Homocysteine<br>(µmol/L) | cotinine | biochemistry | continuous | 2.72E-03 | 7.86E-02 | 8.11E-08 | 1.07E-01 |
| Dietary fiber<br>(gm) | Homocysteine<br>(µmol/L) | food<br>component<br>recall | biochemistry | continuous | 2.18E-04 | -8.38E-02 | 5.53E-04 | -1.21E-01 |
| Folate, RBC<br>(ng/mL RBC) | Homocysteine<br>(μmol/L) | nutrients | biochemistry | continuous | 3.12E-03 | -1.54E-01 | 2.54E-08 | -1.73E-01 |
| Gamma-tocopherol<br>(vitamin E)<br>(μg/dL) | Homocysteine<br>(μmol/L) | nutrients | biochemistry | continuous | 9.08E-04 | 1.02E-01 | 2.23E-04 | 8.94E-02 |
| Iron (mg) | Homocysteine<br>(μmol/L) | food<br>component<br>recall | biochemistry | continuous | 1.05E-04 | -1.13E-01 | 6.14E-05 | -1.33E-01 |
| Niacin (mg) | Homocysteine<br>(μmol/L) | food<br>component<br>recall | biochemistry | continuous | 3.85E-03 | -8.82E-02 | 1.72E-03 | -9.91E-02 |
| Phosphorus<br>(mg) | Homocysteine<br>(μmol/L) | food<br>component<br>recall | biochemistry | continuous | 1.09E-02 | -8.83E-02 | 1.24E-03 | -1.08E-01 |
| Potassium<br>(mg) | Homocysteine<br>(μmol/L) | food<br>component<br>recall | biochemistry | continuous | 1.71E-02 | -6.93E-02 | 5.88E-04 | -1.02E-01 |
| Protein (gm) | Homocysteine<br>(μmol/L) | food<br>component<br>recall | biochemistry | continuous | 3.43E-03 | -8.33E-02 | 9.07E-03 | -1.00E-01 |
| Riboflavin<br>(vitamin B2)<br>(mg) | Homocysteine<br>(μmol/L) | food<br>component<br>recall | biochemistry | continuous | 1.38E-03 | -1.07E-01 | 7.37E-05 | -1.11E-01 |
| Thiamin<br>(vitamin B1)<br>(mg) | Homocysteine<br>(μmol/L) | food<br>component<br>recall | biochemistry | continuous | 1.35E-04 | -9.47E-02 | 9.00E-05 | -1.24E-01 |
| Total<br>supplement<br>count | Homocysteine<br>(μmol/L) | supplement<br>use | biochemistry | continuous | 2.11E-04 | -1.07E-01 | 4.32E-06 | -1.05E-01 |
| Alcohol (gm) | Iron, count<br>(µg/dL) | food<br>component<br>recall | biochemistry | continuous | 3.26E-03 | 8.79E-02 | 6.09E-04 | 9.62E-02 |
| Gamma-tocopherol<br>(vitamin E)<br>(µg/dL) | Iron, count<br>(µg/dL) | nutrients | biochemistry | continuous | 1.23E-03 | -9.01E-02 | 3.59E-05 | -8.92E-02 |
| Retinol<br>(vitamin A)<br>(µg/dL) | Iron, count<br>(µg/dL) | nutrients | biochemistry | continuous | 9.10E-08 | 1.68E-01 | 1.62E-08 | 1.91E-01 |
| Retinol<br>(vitamin A)<br>(µg/dL) | Iron, Frozen<br>Serum (µg/dL) | nutrients | blood | continuous | 3.04E-08 | 1.68E-01 | 2.98E-07 | 2.53E-01 |
| Alpha-tocopherol<br>(vitamin E)<br>(µg/dL) | LDL-cholesterol<br>(mg/dL) | nutrients | biochemistry | continuous | 4.02E-06 | 3.31E-01 | 2.88E-06 | 3.22E-01 |
| Gamma-tocopherol<br>(vitamin E)<br>(µg/dL) | LDL-cholesterol<br>(mg/dL) | nutrients | biochemistry | continuous | 2.35E-03 | 2.27E-01 | 8.79E-05 | 2.29E-01 |
| Cadmium<br>(µg/L) | Lymphocyte<br>number | heavy metals | blood | continuous | 4.43E-04 | 1.28E-01 | 1.87E-06 | 1.31E-01 |
| Cotinine<br>(ng/mL) | Lymphocyte<br>number | cotinine | blood | continuous | 2.22E-07 | 1.78E-01 | 2.68E-08 | 1.57E-01 |
| Current<br>cigarette<br>smoker? | Lymphocyte<br>number | smoking<br>behavior | blood | binary | 4.58E-06 | 4.77E-01 | 1.88E-02 | 2.32E-01 |
| Do you now<br>smoke<br>cigarettes? | Lymphocyte<br>number | smoking<br>behavior | blood | categorical | 1.56E-04 |  | 1.48E-05 |  |
| Does anyone<br>smoke in<br>home? | Lymphocyte<br>number | smoking family | blood | binary | 1.77E-04 | 3.28E-01 | 1.05E-05 | 3.19E-01 |
| Smoked at<br>least 100<br>cigarettes in<br>life | Lymphocyte<br>number | smoking<br>behavior | blood | binary | 2.88E-04 | 2.86E-01 | 3.64E-04 | 2.47E-01 |
| Cadmium<br>(µg/L) | Mean cell<br>hemoglobin (pg) | heavy metals | blood | continuous | 2.30E-04 | 9.86E-02 | 4.29E-05 | 9.52E-02 |
| Cotinine<br>(ng/mL) | Mean cell<br>hemoglobin (pg) | cotinine | blood | continuous | 1.46E-04 | 1.33E-01 | 1.22E-06 | 1.18E-01 |
| Days drink in<br>year | Mean cell<br>hemoglobin (pg) | alcohol use | blood | continuous | 7.28E-06 | 1.59E-01 | 3.08E-03 | 1.19E-01 |
| Do you now<br>smoke<br>cigarettes? | Mean cell<br>hemoglobin (pg) | smoking<br>behavior | blood | categorical | 4.99E-02 |  | 8.19E-05 |  |
| Does anyone<br>smoke in<br>home? | Mean cell<br>hemoglobin (pg) | smoking family | blood | binary | 3.35E-03 | 2.84E-01 | 3.36E-02 | 2.11E-01 |
| Cadmium<br>(µg/L) | Mean cell<br>volume (fL) | heavy metals | blood | continuous | 1.34E-04 | 1.01E-01 | 1.25E-04 | 1.00E-01 |
| Cotinine<br>(ng/mL) | Mean cell<br>volume (fL) | cotinine | blood | continuous | 2.86E-05 | 1.39E-01 | 4.67E-06 | 1.22E-01 |
| Days drink in<br>year | Mean cell<br>volume (fL) | alcohol use | blood | continuous | 1.64E-05 | 1.64E-01 | 9.21E-04 | 1.19E-01 |
| Do you now<br>smoke<br>cigarettes? | Mean cell<br>volume (fL) | smoking<br>behavior | blood | categorical | 1.33E-02 |  | 2.94E-05 |  |
| Does anyone<br>smoke in<br>home? | Mean cell<br>volume (fL) | smoking family | blood | binary | 7.59E-05 | 3.13E-01 | 9.68E-03 | 2.44E-01 |
| Smoked at least 100 cigarettes in life | Mean cell volume (fL) | smoking behavior | blood | binary | 6.37E-03 | 2.37E-01 | 7.62E-09 | 2.50E-01 |
| Cadmium (µg/L) | Monocyte number | heavy metals | blood | continuous | 7.18E-03 | 1.07E-01 | 8.18E-03 | 8.13E-02 |
| Does anyone smoke in home? | Monocyte number | smoking family | blood | binary | 2.25E-04 | 2.81E-01 | 4.10E-02 | 1.91E-01 |
| Folate, RBC (ng/mL RBC) | Red cell count SI | nutrients | blood | continuous | 3.28E-02 | -6.92E-02 | 8.59E-06 | -1.07E-01 |
| Cadmium (µg/L) | Segmented neutrophils number | heavy metals | blood | continuous | 2.92E-03 | 1.19E-01 | 1.61E-05 | 1.35E-01 |
| Cotinine (ng/mL) | Segmented neutrophils number | cotinine | blood | continuous | 1.19E-03 | 1.62E-01 | 7.24E-07 | 1.65E-01 |
| Current cigarette smoker? | Segmented neutrophils number | smoking behavior | blood | binary | 1.88E-05 | 4.37E-01 | 2.85E-04 | 2.25E-01 |
| Do you now smoke cigarettes? | Segmented neutrophils number | smoking behavior | blood | categorical | 1.56E-04 |  | 5.85E-07 |  |
| Does anyone smoke in home? | Segmented neutrophils number | smoking family | blood | binary | 5.15E-04 | 3.35E-01 | 8.40E-07 | 3.25E-01 |
| Smoked at least 100 cigarettes in life | Segmented neutrophils number | smoking behavior | blood | binary | 4.96E-03 | 2.21E-01 | 8.34E-06 | 2.31E-01 |
| Retinol<br>(vitamin A)<br>(µg/dL) | TIBC (µg/dL) | nutrients | biochemistry | continuous | 4.45E-02 | 1.76E-01 | 1.27E-04 | 1.93E-01 |
| Taking birth<br>control | TIBC (µg/dL) | pharmaceutical | biochemistry | binary | 3.67E-05 | 7.00E-01 | 2.64E-07 | 7.87E-01 |
| Retinol<br>(vitamin A)<br>(µg/dL) | Transferrin<br>saturation (%) | nutrients | biochemistry | continuous | 2.69E-03 | 9.55E-02 | 3.79E-04 | 1.69E-01 |
| Alpha-tocopherol<br>(vitamin E)<br>(µg/dL) | Triglycerides<br>(mg/dL) | nutrients | biochemistry | continuous | 1.17E-11 | 4.25E-01 | 8.05E-13 | 4.57E-01 |
| Gamma-tocopherol<br>(vitamin E)<br>(µg/dL) | Triglycerides<br>(mg/dL) | nutrients | biochemistry | continuous | 4.40E-10 | 3.71E-01 | 1.64E-12 | 3.69E-01 |
| Retinol<br>(vitamin A)<br>(µg/dL) | Triglycerides<br>(mg/dL) | nutrients | biochemistry | continuous | 2.22E-10 | 3.06E-01 | 9.30E-12 | 3.07E-01 |
| Retinyl<br>palmitate<br>(µg/dL) | Triglycerides<br>(mg/dL) | nutrients | biochemistry | continuous | 3.24E-05 | 2.73E-01 | 1.20E-05 | 3.27E-01 |
| Magnesium<br>(mg) | Uric acid (mg/dL) | food<br>component<br>recall | biochemistry | continuous | 4.98E-02 | -6.17E-02 | 3.50E-02 | -5.90E-02 |
| Phosphorus<br>(mg) | Uric acid (mg/dL) | food<br>component<br>recall | biochemistry | continuous | 2.24E-02 | -7.27E-02 | 4.14E-02 | -6.71E-02 |
| Retinol<br>(vitamin A)<br>(µg/dL) | Uric acid (mg/dL) | nutrients | biochemistry | continuous | 1.71E-08 | 2.08E-01 | 2.12E-10 | 2.30E-01 |
| Riboflavin<br>(vitamin B2)<br>(mg) | Uric acid (mg/dL) | food<br>component<br>recall | biochemistry | continuous | 8.16E-04 | -8.02E-02 | 2.77E-04 | -8.25E-02 |

Replicated environment-phenotype associations with their discovery and replication Bonferroni-adjusted p-values, categories, data types, and effect sizes.

### Exposures with cross-phenotype associations

To determine which exposures were involved with multiple phenotypes, we evaluated the top 106 exposure-phenotype associations with Bonferroni-significant results in discovery and replication datasets for exposures with cross-phenotype associations (i.e., exposures that demonstrated significant associations with more than one phenotype). We identified 19 exposures with cross-phenotype associations. These exposures belonged to the categories of vitamins (6), smoking (5), dietary supplements (nonmetals and metals) (4), metal pollutants (2), drugs and chemicals (1), and alcohol (1) (Fig. 2). The smoking exposure category had the highest number of associations (34), including cotinine (11) and survey questions: “Does anyone smoke in home?” (9), “Do you now smoke cigarettes?” (6), “Current cigarette smoker?” (4), and “Smoked at least 100 cigarettes in life” (3). The individual exposures that had the most cross-phenotype associations were retinol (14), cotinine (11), cadmium (10), “Does anyone smoke in home?” (9), and gamma-tocopherol (9) (Table 1, Supplemental Table 1, Fig. 3).

**Fig. 3.**
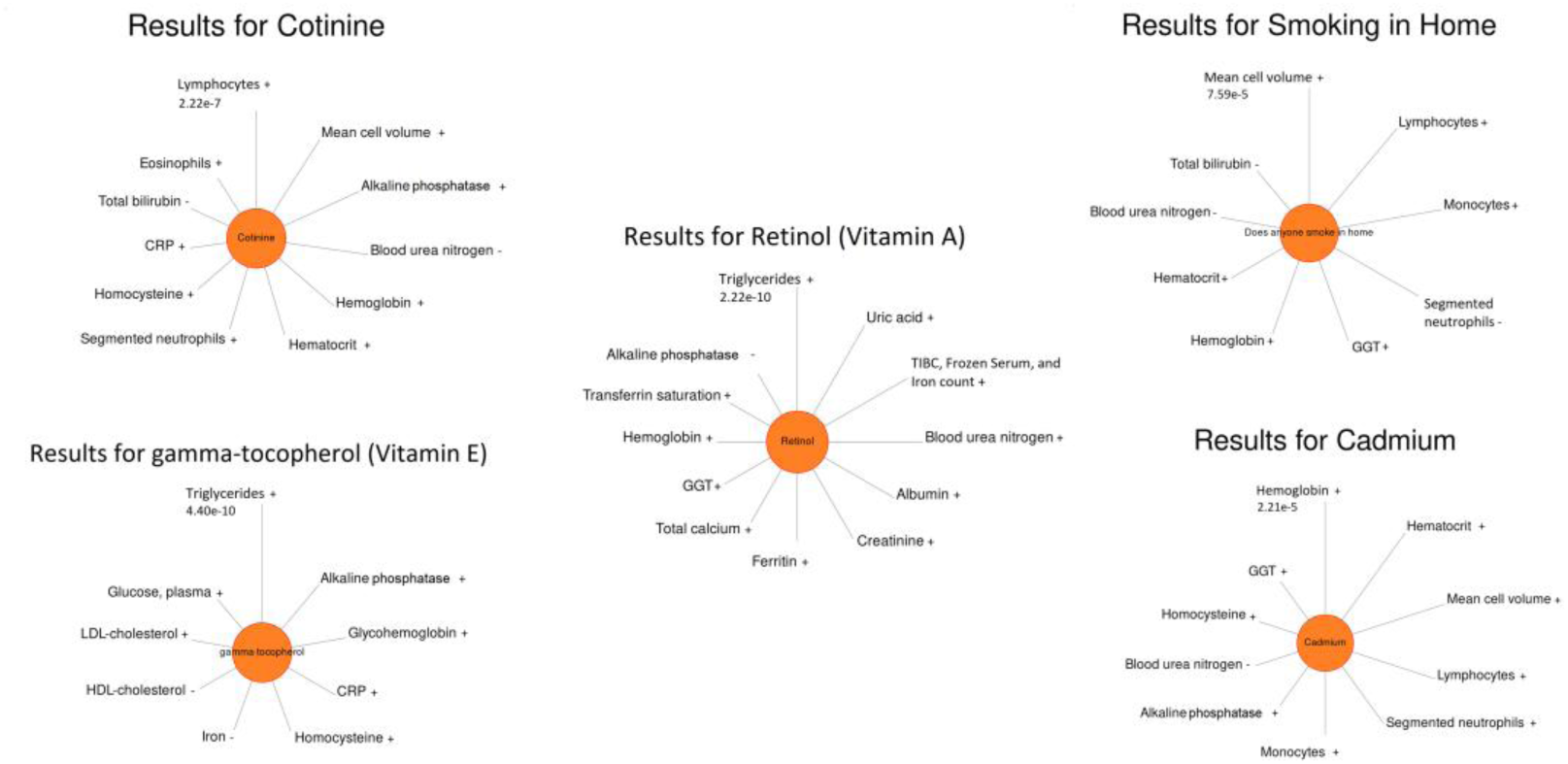
Exposures with multiple cross-phenotype associations. Sun plot of results for exposures retinol, cotinine, cadmium, *Does anyone smoke in home?* and gamma-tocopherol. Replicated cross-phenotype associations plotted clockwise with the smallest p-value at the top. These exposures had the largest number of cross-phenotypic effects. The length of the line corresponds to the −log(p-value) for each association, and the longest line shows the most significant result and p-value. At the end of the lines, the positive (+) and negative (−) indicate the direction of the effect of each association. The sun plots were generated using PheWAS-View70.

Retinol had the largest number of cross-phenotype associations, which included associations with triglycerides (P_d_ = 1.24 × 10^-14^, β_d_ = 0.31; Pᵣ = 4.85 × 10^-15^, βᵣ = 0.31), uric acid (P_d_ = 9.57 × 10^-13^, β_d_ = 0.21; Pᵣ = 1.11 × 10^-13^, βᵣ = 0.23), frozen serum iron (P_d_ = 1.70 × 10^-12^, β_d_ = 0.17; Pᵣ = 1.55 × 10^-10^, βᵣ = 0.25), blood urea nitrogen (P_d_ = 8.04 × 10^-12^, β_d_ = 0.25; Pᵣ = 5.47 × 10^-13^, βᵣ = 0.24), ferritin (P_d_ = 6.91 × 10^-9^, β_d_ = 0.14; Pᵣ = 8.41 × 10^-11^, βᵣ = 0.28), gamma-glutamyl transferase (GGT) (P_d_ = 6.91 × 10^-9^, β_d_ = 0.18; Pᵣ = 1.37 × 10^-12^, βᵣ = 0.19), hemoglobin (P_d_ = 2.48 × 10^-8^, β_d_ = 0.10; Pᵣ = 7.39 × 10^-6^, βᵣ = 0.09), and total iron-binding capacity (TIBC) (P_d_ = 2.48 × 10^-6^, β_d_ = 0.18; Pᵣ = 6.64 × 10^-8^, βᵣ = 0.24).

Cotinine, a biomarker for smoking exposure, had cross-phenotype associations, among which nine had positive beta values. These included WBC phenotypes, such as lymphocyte number (P_d_ = 1.24 × 10^-11^, β_d_ = 0.18; Pᵣ = 1.40 × 10^-11^, βᵣ = 0.16), segmented neutrophil number (P_d_ = 6.65 × 10^-8^, β_d_ = 0.16; Pᵣ = 3.78 × 10^-10^, βᵣ = 0.17), and eosinophils (P_d_ = 2.47 × 10^-6^, β_d_ = 0.07; Pᵣ = 2.61 × 10^-7^, β_r_= 0.08). Cotinine also had positive associations with RBC measurements, including mean cell volume (P_d_ = 1.60 × 10^-9^, β_d_ = 0.14; Pᵣ = 2.43 × 10^-9^, βᵣ = 0.24), hemoglobin (P_d_ = 1.76 × 10^-8^, β_d_ = 0.11; Pᵣ = 8.04 × 10^-12^, βᵣ = 0.12), and hematocrit (P_d_ = 2.45 × 10^-8^, β_d_ = 0.10; Pᵣ = 2.38 × 10^-11^, β_r_ = 0.11).

Cotinine demonstrated two cross-phenotype associations with negative beta values: blood urea nitrogen (P_d_ = 3.41 × 10^-9^, β_d_ = -0.12; Pᵣ = 3.12 × 10^-11^, βᵣ = −0.12) and bilirubin (P_d_ = 1.29 × 10^-6^, β_d_ = −0.11; Pᵣ = 1.37 × 10^-6^, βᵣ = -0.09).

The survey question, “Does anyone smoke in the home?” labelled as “Smoking in home” was significantly associated with a number of WBC and RBC traits with positive beta values. The WBC count phenotypes included lymphocytes (P_d_ = 8.71 × 10^-9^, β_d_ = 0.33; Pᵣ = 7.69 × 10^-9^, βᵣ = 0.32), monocytes (P_d_ = 1.25 × 10^-8^, β_d_ = 0.28; Pᵣ = 2.14 × 10^-5^, βᵣ = 0.19), and segmented neutrophils (P_d_ = 2.87 × 10^-8^, β_d_ = 0.34; Pᵣ = 4.38 × 10^-10^, βᵣ = 0.33). Additionally, they were associated with RBC measurements, including mean cell volume (P_d_ = 4.24 × 10^-9^, β_d_ = 0.31; Pᵣ = 5.05 × 10^-6^, βᵣ = 0.24) and hematocrit (P_d_ = 5.60 × 10^-9^, β_d_ = 0.21; Pᵣ = 6.37 × 10^-8^, βᵣ = 0.23)

Cadmium demonstrated 11 cross-phenotype associations. The top cross-phenotype associations were related to blood measurements, including hematocrit (P_d_ = 2.22 × 10^-9^, β_d_ = 0.12; Pᵣ = 2.93 × 10^-11^, βᵣ = 0.13), mean cell volume (P_d_ = 7.48 × 10^-9^, β_d_ = 0.10; Pᵣ = 6.52 × 10^-8^, βᵣ = 0.10), and lymphocyte number (P_d_ = 2.47 × 10^-8^, β_d_ = 0.13; Pᵣ = 9.74 × 10^-10^, βᵣ = 0.13).

Gamma-tocopherol exhibited nine significant cross-phenotype associations, some of which overlapped with alpha-tocopherol, showing positive beta values with lipid-related phenotypes such as triglycerides (alpha-tocopherol: P_d_ = 6.51 × 10^-16^, β_d_ = 0.42; Pᵣ = 4.20 × 10^-16^, βᵣ = 0.46; gamma-tocopherol: P_d_ = 2.46 × 10^-14^, β_d_ = 0.37; Pᵣ = 8.54 × 10^-16^, βᵣ = 0.37) and LDL cholesterol (alpha-tocopherol: P_d_ = 2.24 × 10^-10^, β_d_ = 0.33; Pᵣ = 1.50 × 10^-9^, βᵣ = 0.32; gamma-tocopherol: P_d_ = 1.31 × 10^-7^, β_d_ = 0.23; Pᵣ = 4.58 × 10^-8^, βᵣ = 0.23). While alpha-tocopherol was associated with a negative beta value with homocysteine (P_d_ = 1.04 × 10^-7^, β_d_ = −0.10; Pᵣ = 1.61 × 10^-9^, βᵣ = -0.11), the association between gamma-tocopherol and homocysteine showed a positive beta value (P_d_ = 5.07 × 10^-8^, β_d_ = 0.09; Pᵣ = 1.16 × 10^-7^, βᵣ = 0.09).

### Phenotypes with cross-exposure associations

Similar to identifying exposures with cross-phenotype associations, we determined which phenotypes were involved with multiple exposures. As such, we evaluated the top 106 exposure-phenotype associations with Bonferroni-significant results in discovery and replication datasets for phenotypes with cross-exposure associations (i.e., phenotypes that demonstrated significant associations with more than one exposure). We identified 23 phenotypes with cross-exposure associations, classified as metabolic products (4), RBC measurements (4), WBCs (3), iron measurements (3), lipids (3), blood proteins (3) enzymes (2), and amino acids (1) (Table 1 & Fig. 2). When we considered associations within each phenotype category, phenotypes from the metabolic products category had the most associations (17), which included the traits blood urea nitrogen (7), uric acid (4), creatinine (3), and total bilirubin (3). At the individual level, phenotypes with the most cross-exposure associations were homocysteine (14), blood urea nitrogen (7), lymphocyte number (6), segmented neutrophil number (6), and mean cell volume (6) (Table 1, Supplemental Table 1, Fig. 4).

**Fig. 4.**
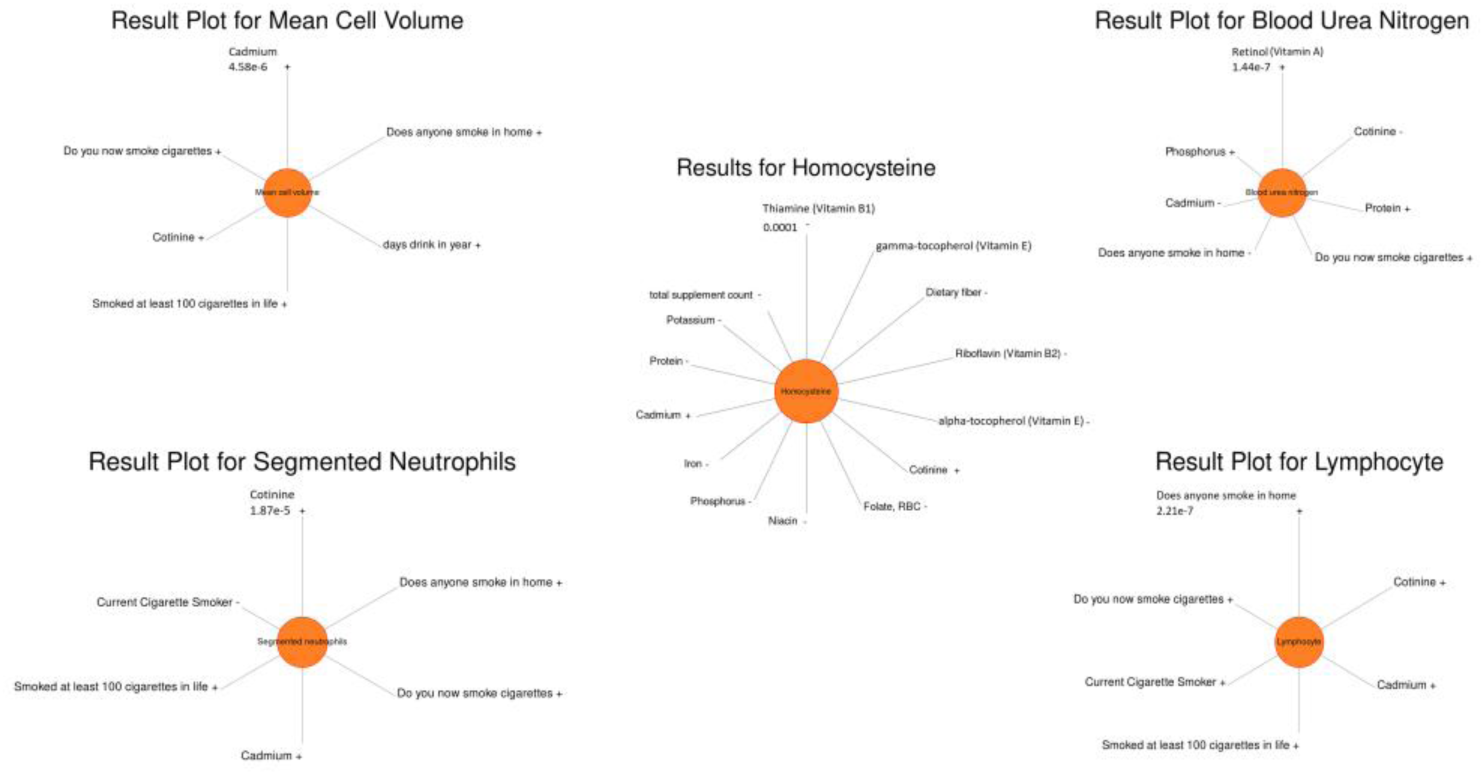
Phenotypes with multiple cross-exposure associations. Sun plot of results for phenotypes homocysteine, lymphocyte count, segmented neutrophil count, blood urea nitrogen, and mean cell volume. Significant cross-exposure associations are plotted clockwise, with the smallest p-value at the top. These phenotypes exhibited the greatest cross-exposure connections. The length of the line corresponds to the −log(p-value) for each association, and the longest line shows the most significant result and p-value. At the end of the lines, the positive (+) and negative (−) indicate the direction of the effect of each association. The sun plots were generated using PheWAS-View70.

Homocysteine associated with multiple vitamin exposures with a negative beta coefficient. For instance, homocysteine was associated with folate RBC (vitamin B9) (P_d_ = 1.74 × 10^-7^, β_d_ = −0.15; Pᵣ = 1.32 × 10^-11^, βᵣ = −0.17), riboflavin (vitamin B2) (P_d_ = 7.73 × 10^-8^, β_d_ = −0.11; Pᵣ = 3.84 × 10^-8^, βᵣ = −0.11), total supplement count (P_d_ = 1.18 × 10^-8^, β_d_ = −0.11; Pᵣ = 2.25 × 10^-9^, βᵣ = −0.11), potassium (P_d_ = 9.52 × 10^-7^, β_d_ = −0.07; Pᵣ = 3.06 × 10^-7^, βᵣ = −0.10), alpha-tocopherol (P_d_ = 1.03 × 10^-7^, β_d_ = −0.09; Pᵣ = 1.61 × 10^-9^, βᵣ = −0.11) and phosphorus (P_d_ = 6.07 × 10^-7^, β_d_ = −0.09; Pᵣ = 6.46 × 10^-7^, βᵣ = −0.11). Homocysteine was associated with gamma-tocopherols (P_d_ = 5.07 × 10^-8^, β_d_ = 0.10; Pᵣ = 1.16 × 10^-7^, βᵣ = 0.09) with positive beta values.

Lymphocyte and segmented neutrophil counts were associated with cotinine and cadmium, as previously described, and were also associated with other smoking-related exposures. Furthermore, lymphocyte levels were associated with survey questions “Current cigarette smoker?” (P_d_ = 2.56 × 10^-10,^ β_d_ = 0.48; Pᵣ = 9.81 × 10^-6^, βᵣ = 0.23), “Do you now smoke cigarettes?” (P_d_ = 8.71 × 10^-9^; Pᵣ = 7.69 × 10^-9^), and “Smoked at least 100 cigarettes in life” (P_d_ = 1.61 × 10^-8^, β_d_ = 0.29; Pᵣ = 1.89 × 10^-7^, βᵣ = 0.25). Similarly, segmented neutrophil count was associated with “Current cigarette smoker?” (P_d_ = 1.88 × 10^-5^, β_d_ = 0.44; Pᵣ = 1.48 × 10^-7^, βᵣ = 0.23), “Do you now smoke cigarettes?” (P_d_ = 1.56 × 10^-4^; Pᵣ = 3.05 × 10^-10^), and “Smoked at least 100 cigarettes in life” (P_d_ = 2.77 × 10^-7^, β_d_ = 0.22; Pᵣ = 4.35 × 10^-9^, βᵣ = 0.23).

Finally, mean cell volume demonstrated the largest number of cross-exposure of significant associations with exposures in the smoking category. The top cross-exposure associations for this phenotype related to smoking: “Do you now smoke cigarettes?” (P_d_ = 7.44 × 10^-7^; Pᵣ = 1.53 × 10^-8^) and “Smoked at least 100 cigarettes in life” (P_d_ = 3.56 × 10^-7^, β_d_ = 0.24; Pᵣ = 3.97 × 10^-12^, βᵣ = 0.25).

### Interconnections between the phenome and exposome

To visualize the intricate interplay among the 106 exposure-phenotype associations with Bonferroni-significance across the discovery and replication datasets, we generated a network map that illustrates the complexity of results across the phenome and exposome (Fig. 2). Among the 106 significant associations, 69 exhibited positive beta values, indicating relationships between exposures and phenotypes, whereas 29 demonstrated negative beta values, suggesting inverse associations. The remaining six associations involved categorical predictors, so no beta coefficients were reported because there was no single effect estimate [see Methods]. The resulting network reveals cross-phenotype and cross-exposure links, with some variables acting as highly connected nodes due to their associations with multiple traits. Retinol (Vitamin A) had the highest number of statistically significant cross-phenotype associations, highlighting its extensive links across diverse biological measures.

The visual layout of the network reveals distinct clusters of interconnected variables, where certain phenotypes and exposures act as bridges linking different variables. The size of each node represents its degree centrality, with larger nodes indicating variables that have a higher number of significant connections. Nodes such as retinol (vitamin A), homocysteine, cotinine, and cadmium appear larger, reflecting their roles as key points of interaction within the system. Their positioning sets them as central hubs within the network. Conversely, exposures and phenotypes with fewer connections form more isolated nodes on the periphery. Variables with only one connection, such as niacin (mg) or thiamin (vitamin B1) (mg), appear as smaller nodes, resembling appendages extending from the main network. These periphery nodes, while statistically significant, exhibit more limited influence in terms of cross-exposure or cross-phenotype associations within this study. This structural organization provides insight into how different exposures and biological markers are related, illustrating systemic interdependencies and hierarchical relationships. The varying node sizes and connection densities help to identify key variables, clusters, and association patterns.

### PheEWAS results stratified by race-ethnicity

In addition to the multi-ethnic PheEWAS described above, we performed a PheEWAS stratified by self-identified Non-Hispanic Blacks (NHB), Non-Hispanic Whites (NHW), and Mexican Americans (MA) race/ethnicity using the same QC and analytical approaches as our unstratified PheEWAS, as described in the Methods section. For each stratified PheEWAS, in the discovery datasets NHB (1,335), NHW (1,887), and MA (1,887) had FDR < 0.1 and were considered for the replication datasets, which NHB (276), NHW (194), and MA (945) had the same direction of effect with FDR < 0.1. When comparing across the discovery and replication datasets, there were 106 significant exposure-phenotype results in total that had a p-value < 0.05 (in both) with Bonferroni adjustment for all three groups: NHB (7), NHW (88), and MA (11). Notably, the total number of significant associations in the stratified PheEWAS, equal to that of the unstratified PheEWAS, is purely coincidental, as stratification altered the composition and distribution of associations (Table 2, Fig. 5). Ninety-five of the 106 associations from the unstratified analysis were identified in at least one stratum. The top five exposure-phenotype associations (retinol with blood urea nitrogen, gamma-tocopherol with triglycerides, alpha-tocopherol with triglycerides, retinol with triglycerides, and retinol with uric acid) were significant in all three strata and in the unstratified PheEWAS (Table 2, Fig. 6). Two associations were shared between NHB and NHW (retinol with creatinine and “smoked at least 100 cigarettes in life” with lymphocyte number), four associations were shared between the MA and NHW strata (alpha-tocopherol with LDL cholesterol, retinol with ferritin, retinol with total calcium, and thallium with creatinine) and no associations were shared between the NHB and MA strata (Fig 6, Supplemental Table 2). Eleven associations were unique to the race/ethnicity-specific analysis, two of which were unique to the MA stratum and nine to the NHW stratum (Table 2 & Supplemental Table 2). The two unique associations from the MA stratum were urinary cadmium with urinary creatinine (Bonferroni-adjusted P_d_ = 7.83 × 10^−4^, β_d_ =1.27; Bonferroni-adjusted P_r_ = 8.77 × 10^−5^, β_r_ = 0.66) and platinum with eosinophils (Bonferroni-adjusted P_d_ = 3.12 × 10^−2^, β_d_ =−35.01; Bonferroni-adjusted P_r_ = 3.89 × 10^−4^, β_r_ = −2.72). Two examples from the NHW stratum include an association with a positive beta coefficient, between lead in urine and creatinine in urine (Bonferroni-adjusted P_d_ = 6.57 × 10^−4^, β_d_ =0.72; Bonferroni-adjusted P_r_ = 2.36 × 10^−10^, β_r_ =1.14), and an association with a negative beta coefficient between trans-beta-carotene and homocysteine (Bonferroni-adjusted P_d_ = 3.31 × 10^−2^, β_d_ =−0.10; Bonferroni-adjusted P_r_ = 6.55 × 10^−6^, β_r_ = −0.12).

**Fig. 5.**
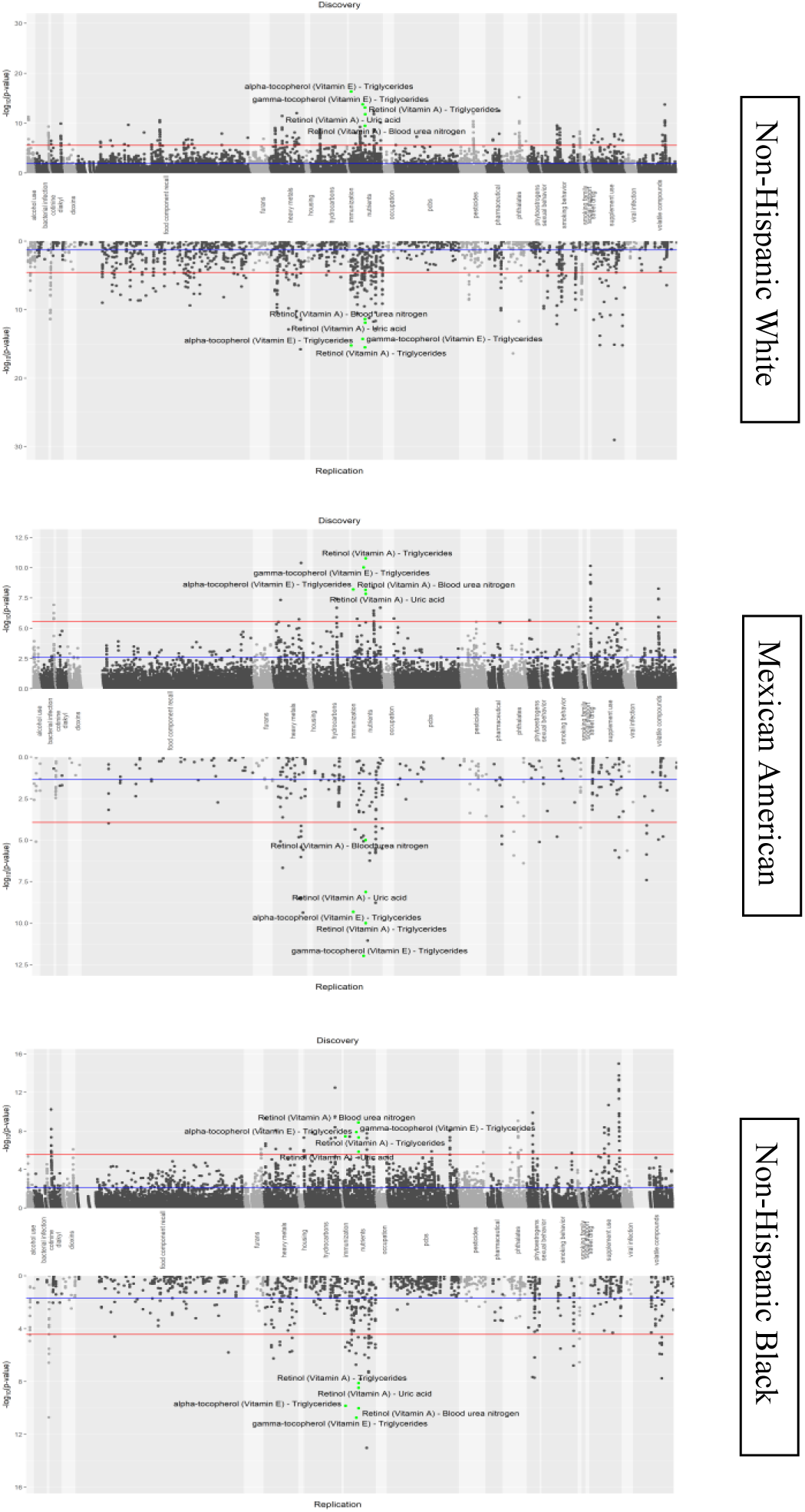
Stratified PheEWAS results. The three Hudson plots are arranged by race/ethnicity strata: Non-Hispanic White, Mexican American, and Non-Hispanic Black. On the x-axis are exposure-phenotype associations, ordered by exposure category, and on the y-axis is the −log10(p-value). The red lines denote a Bonferroni-adjusted p-value of 0.05, and the blue lines denote the threshold for FDR of 0.1. The top replicating results in the nutrients category that are significant after correction are labeled. Hudson plots were created using an R package available at https://github.com/anastasia-lucas/hudson67

**Fig. 6.**
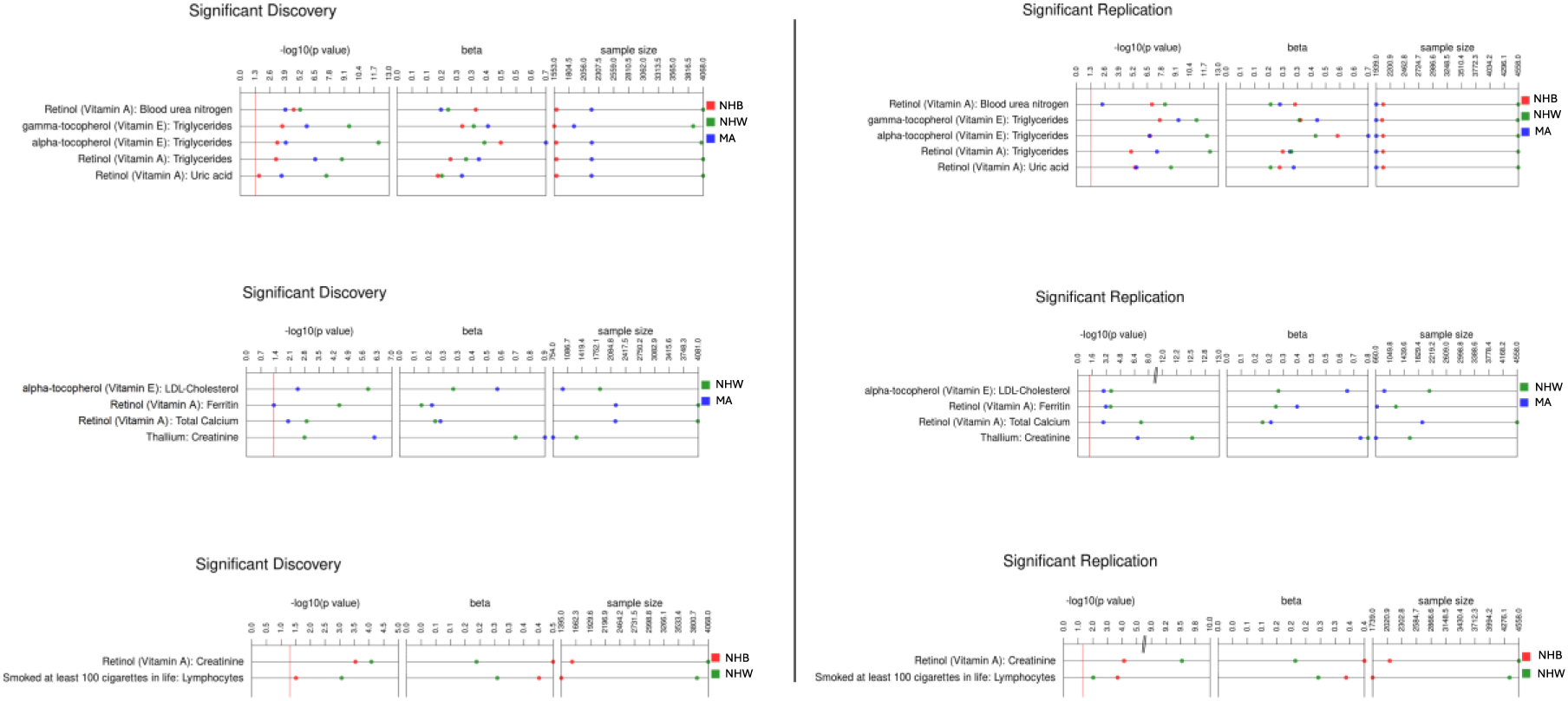
Significant stratified PheEWAS results. This plot depicts exposure-phenotype associations with Bonferroni-adjusted p-values < 0.05 that overlap for each race-ethnicity stratum. Each color represents a stratified race-ethnicity stratum, red for Non-Hispanic Black (NHB), green for Non-Hispanic White (NHW), and blue for Mexican American (MA), as represented in the NHANES data displaying the same direction of effect. Presented also are the Bonferroni-adjusted p-value for each association and the comparison between all the beta values, indicating the magnitude of effect and varying sample sizes70. *Defined abbreviations found in the tables: Gamma-glutamyl transferase (GGT), Red blood cell (RBC), Low-density lipoprotein (LDL), High-density lipoprotein (HDL), Total iron binding capacity (TIBC)

**Table 2:**
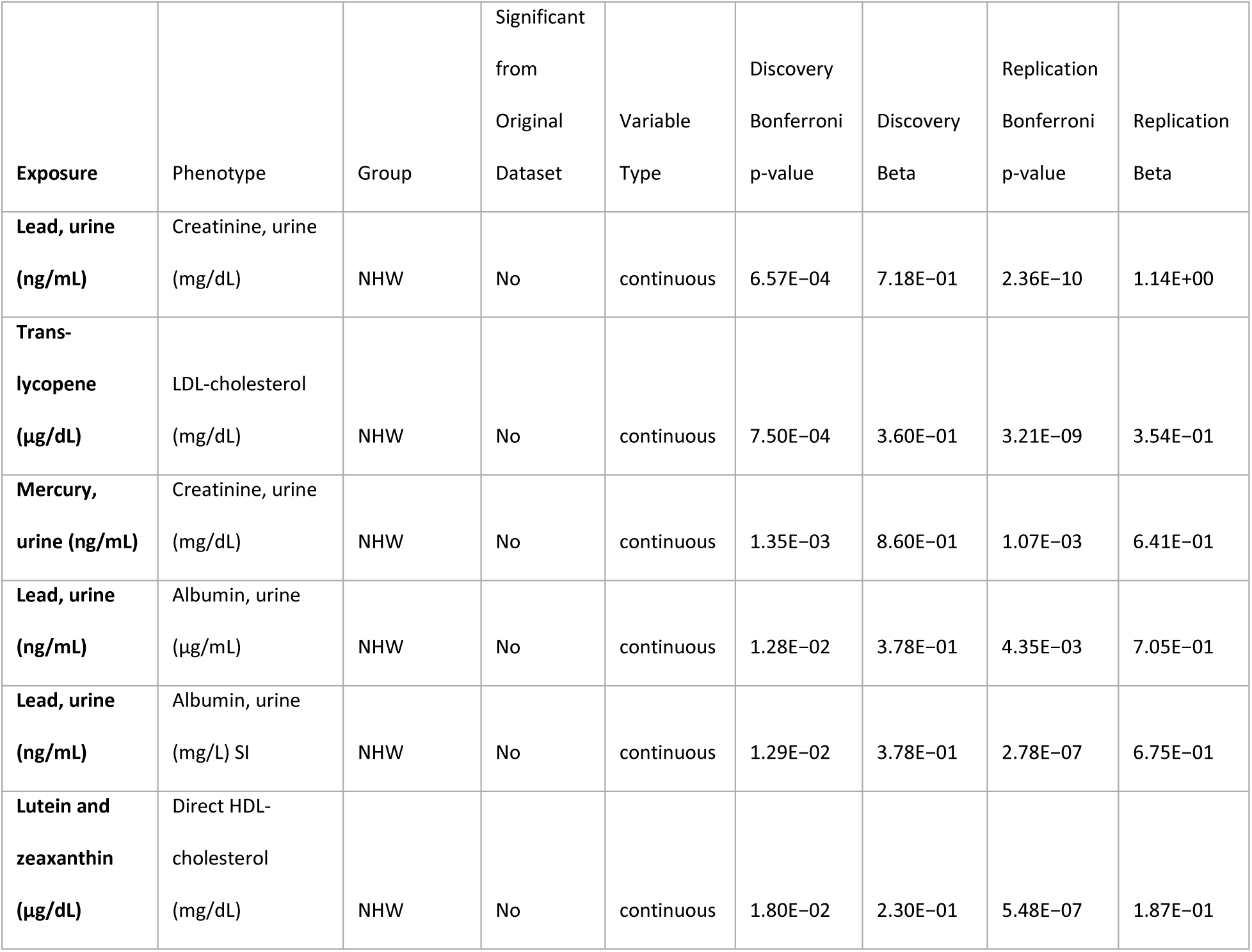
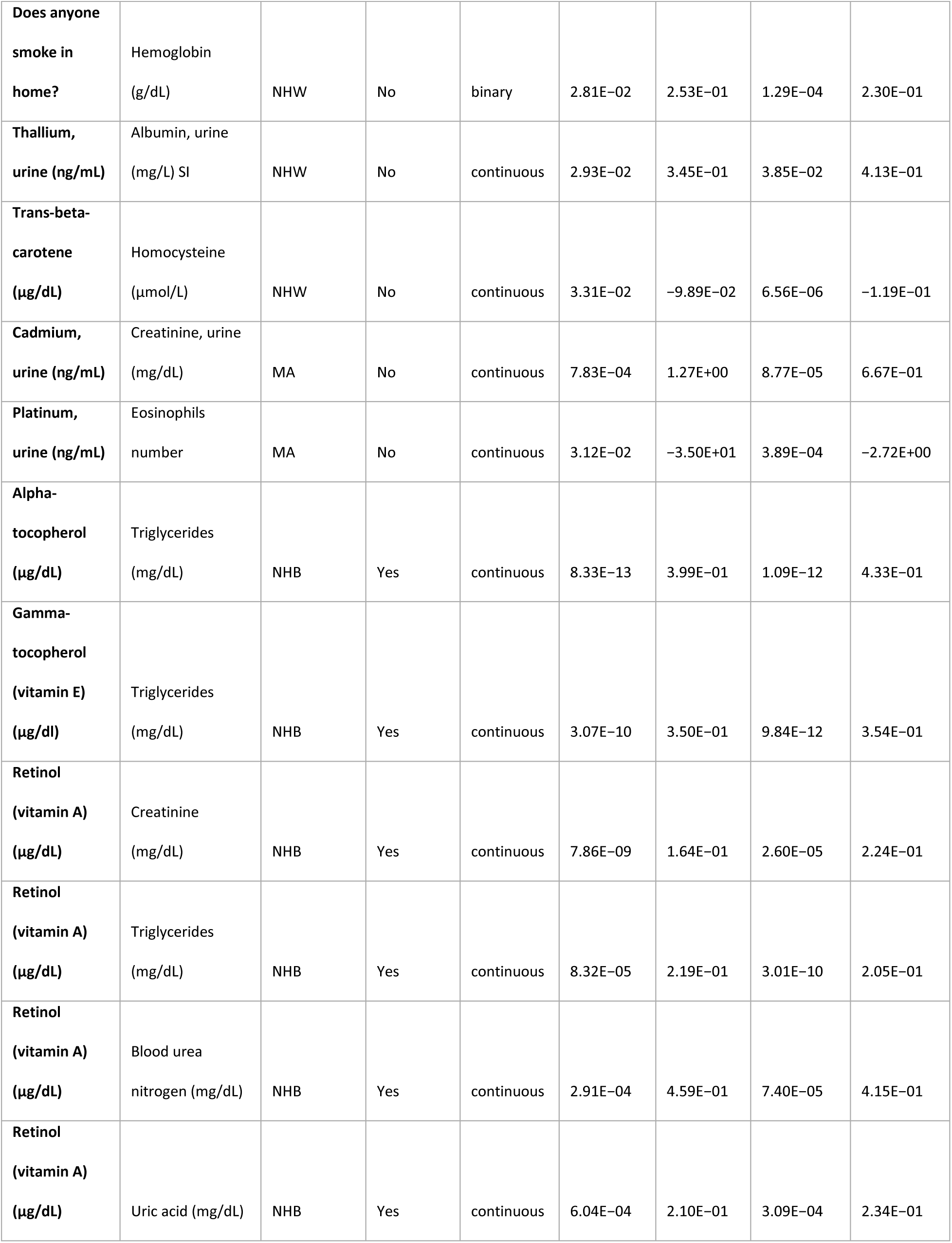
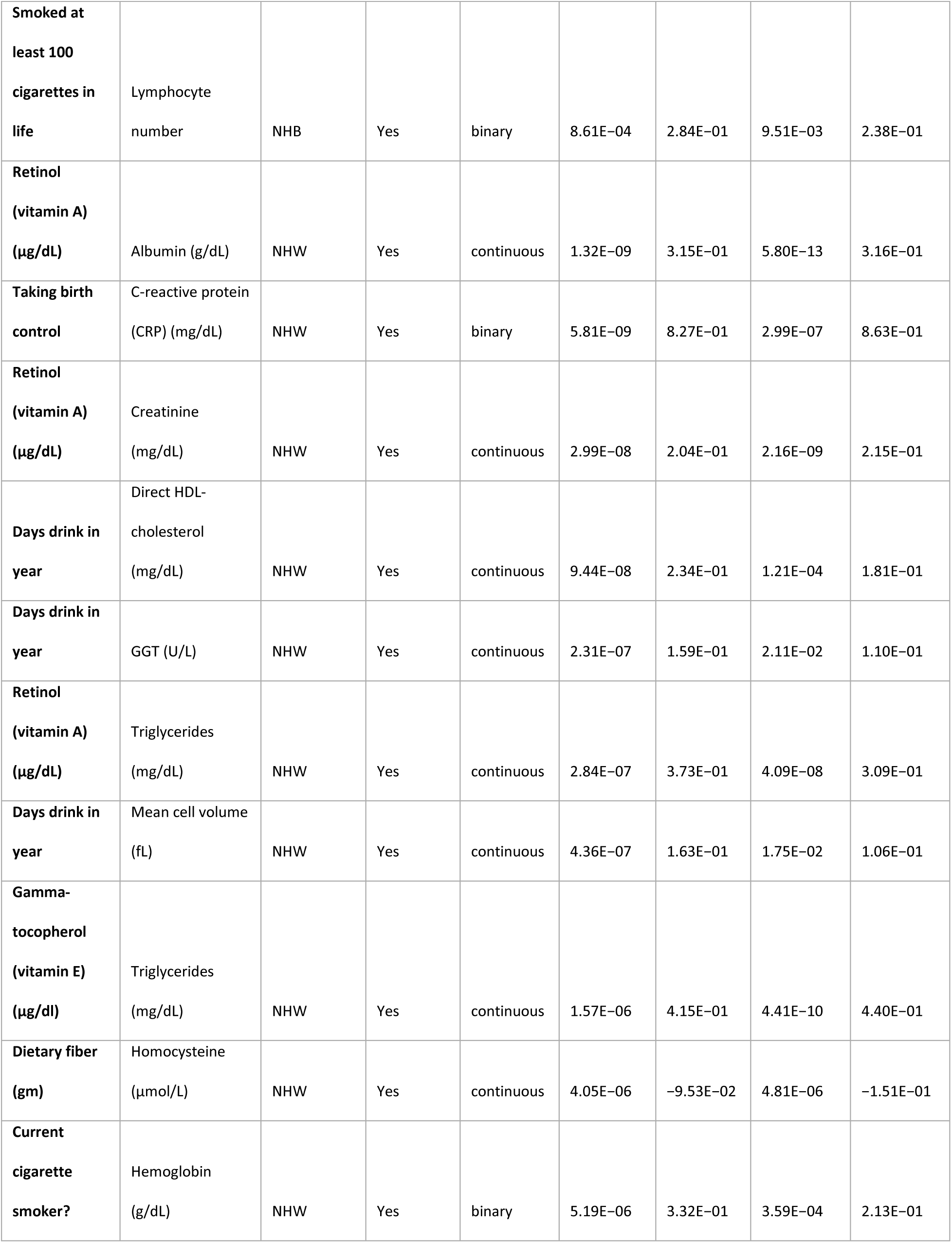
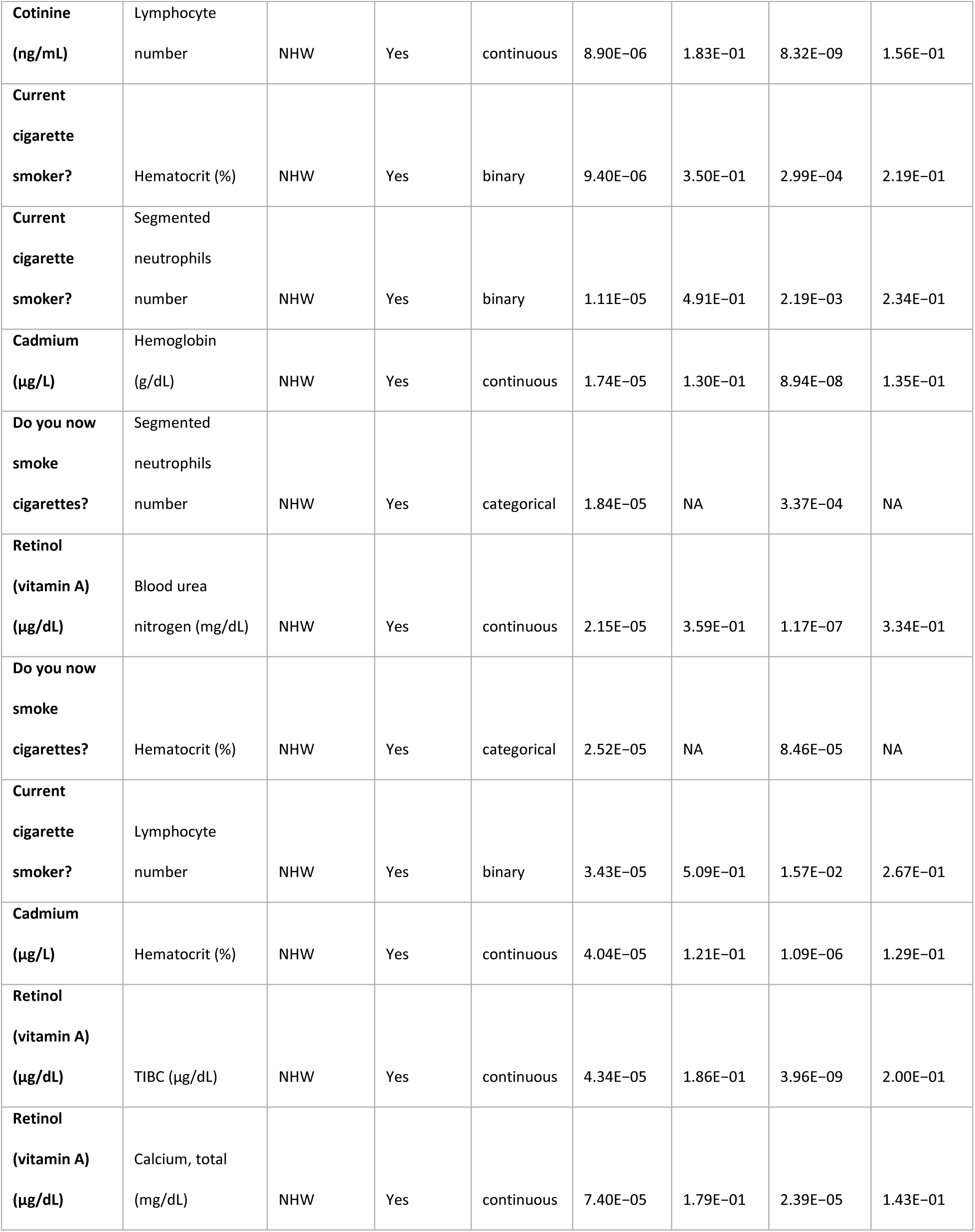
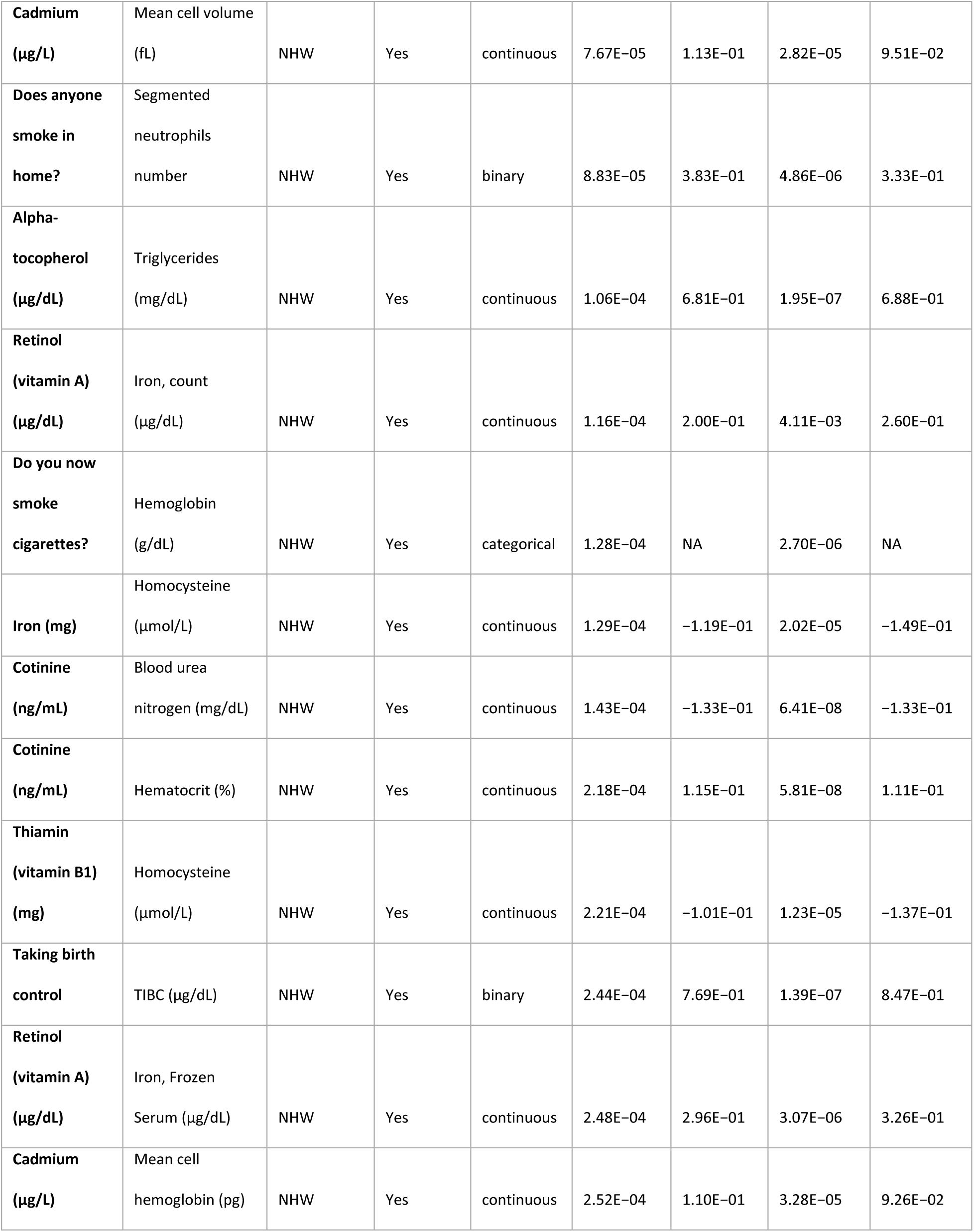
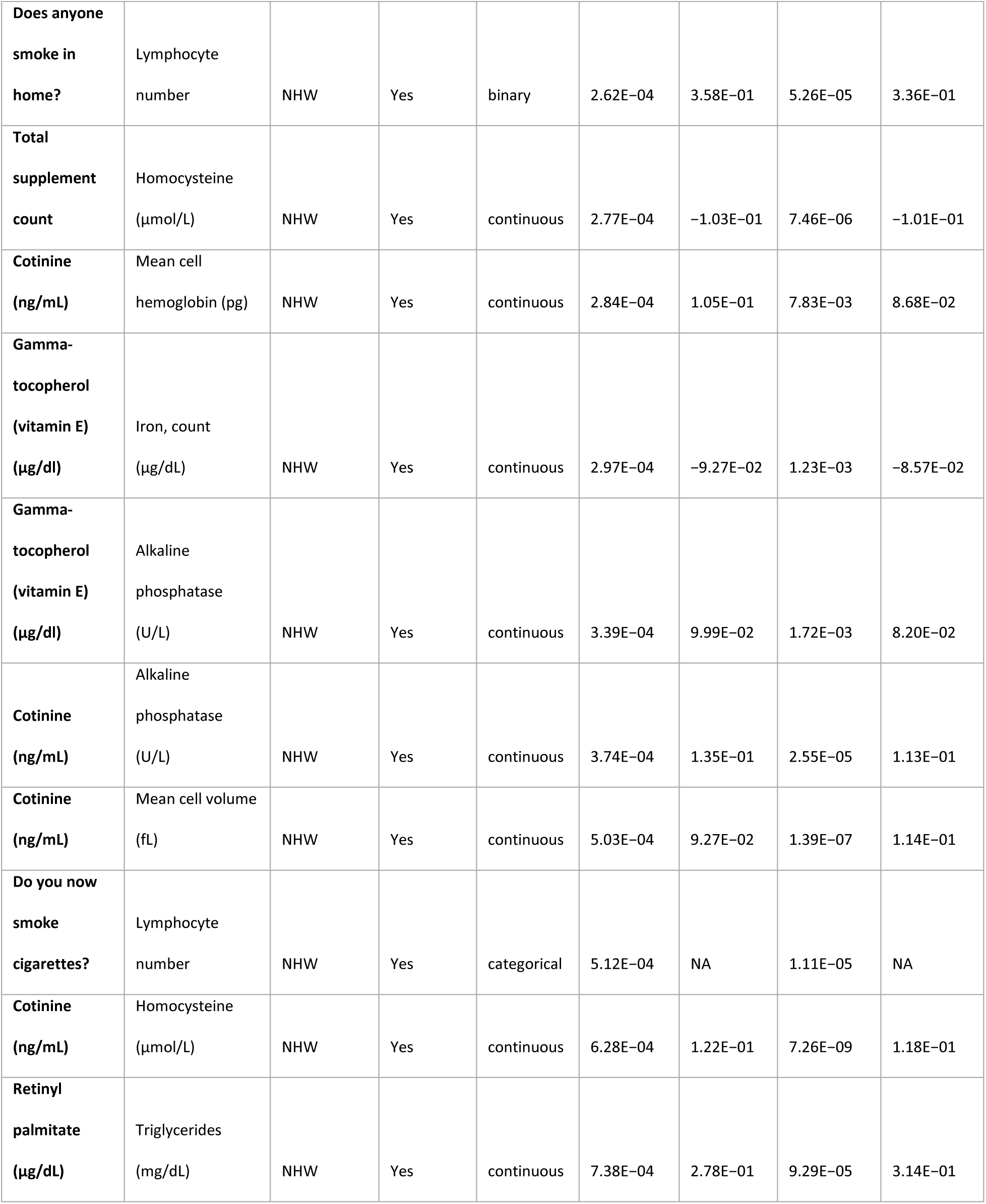
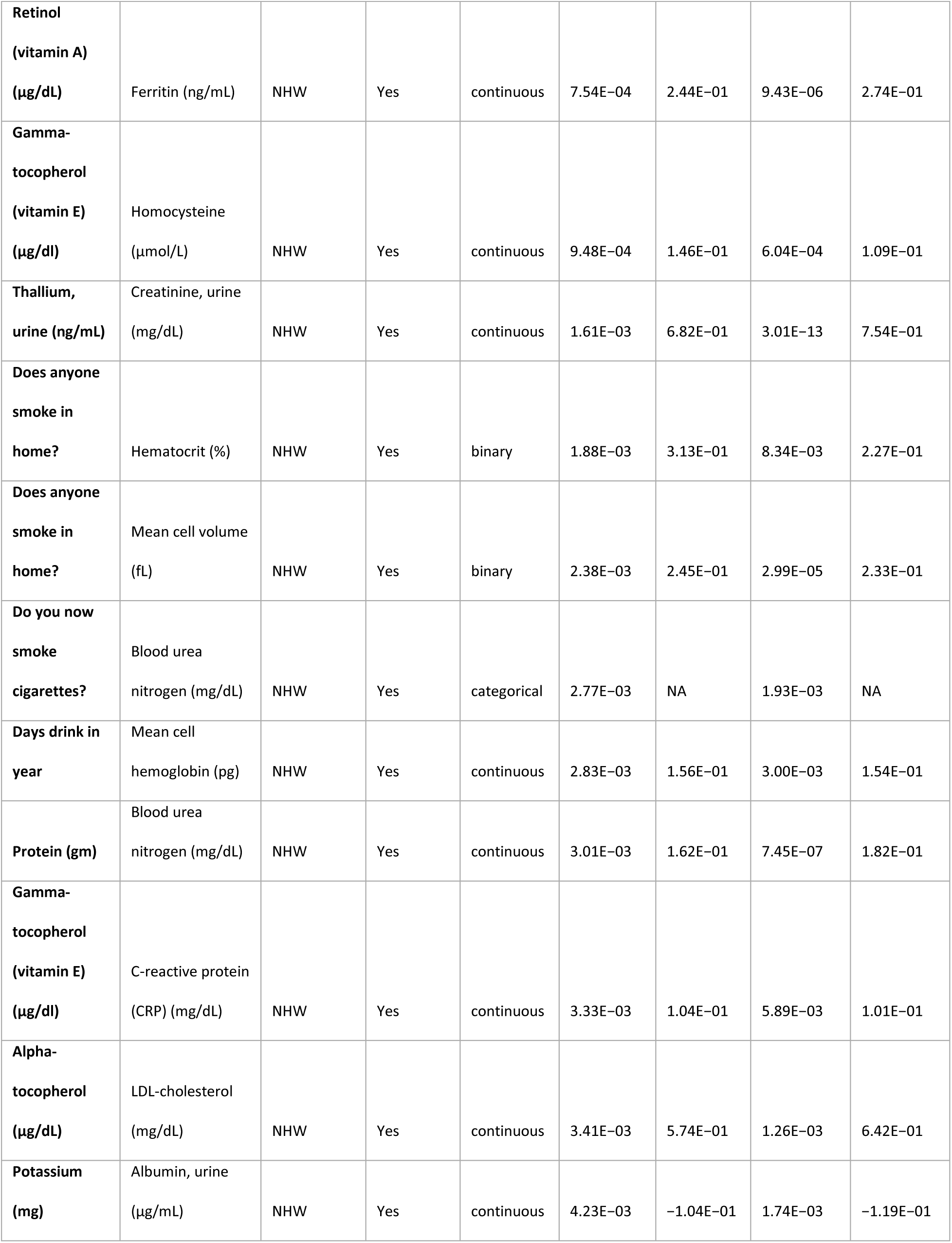
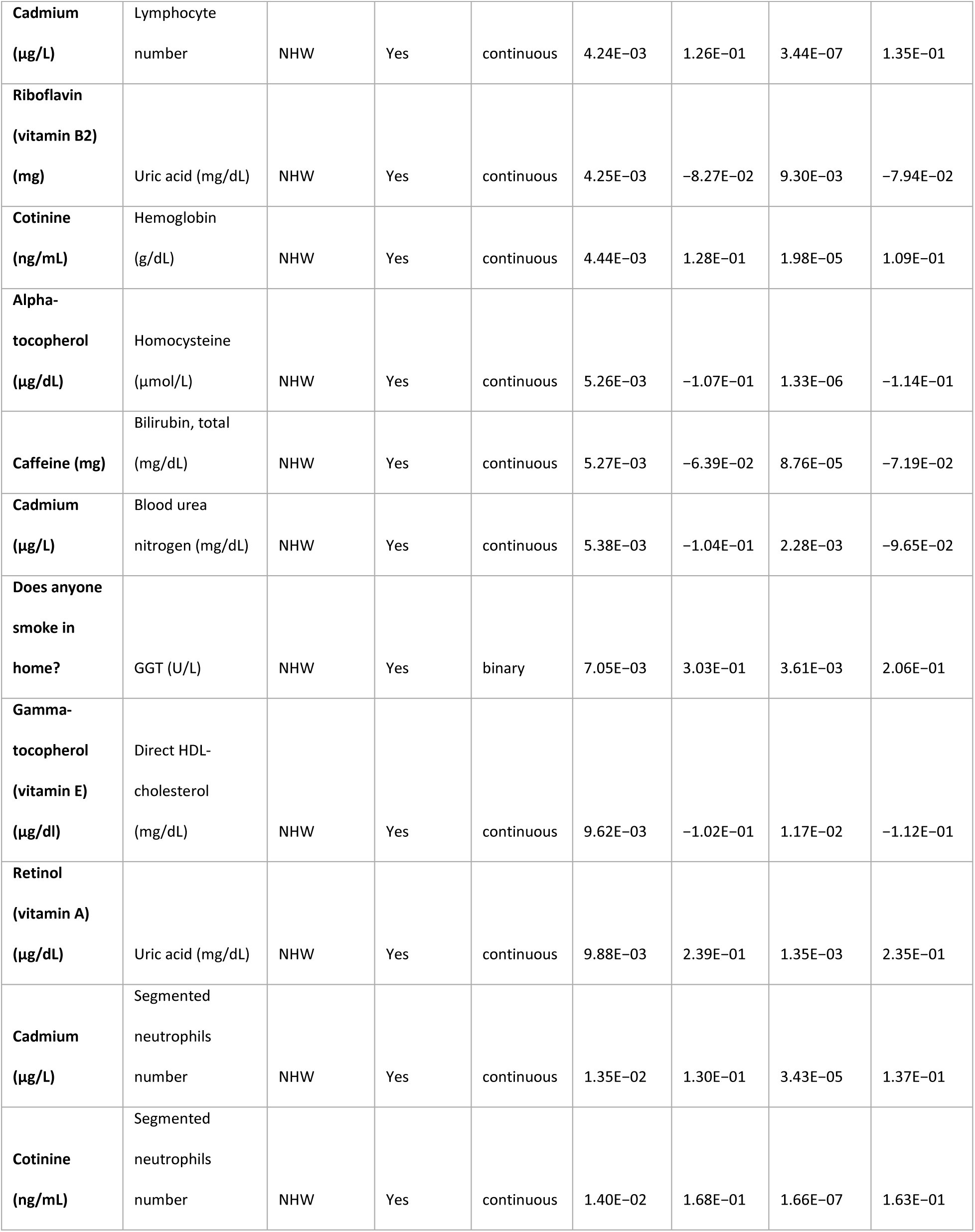
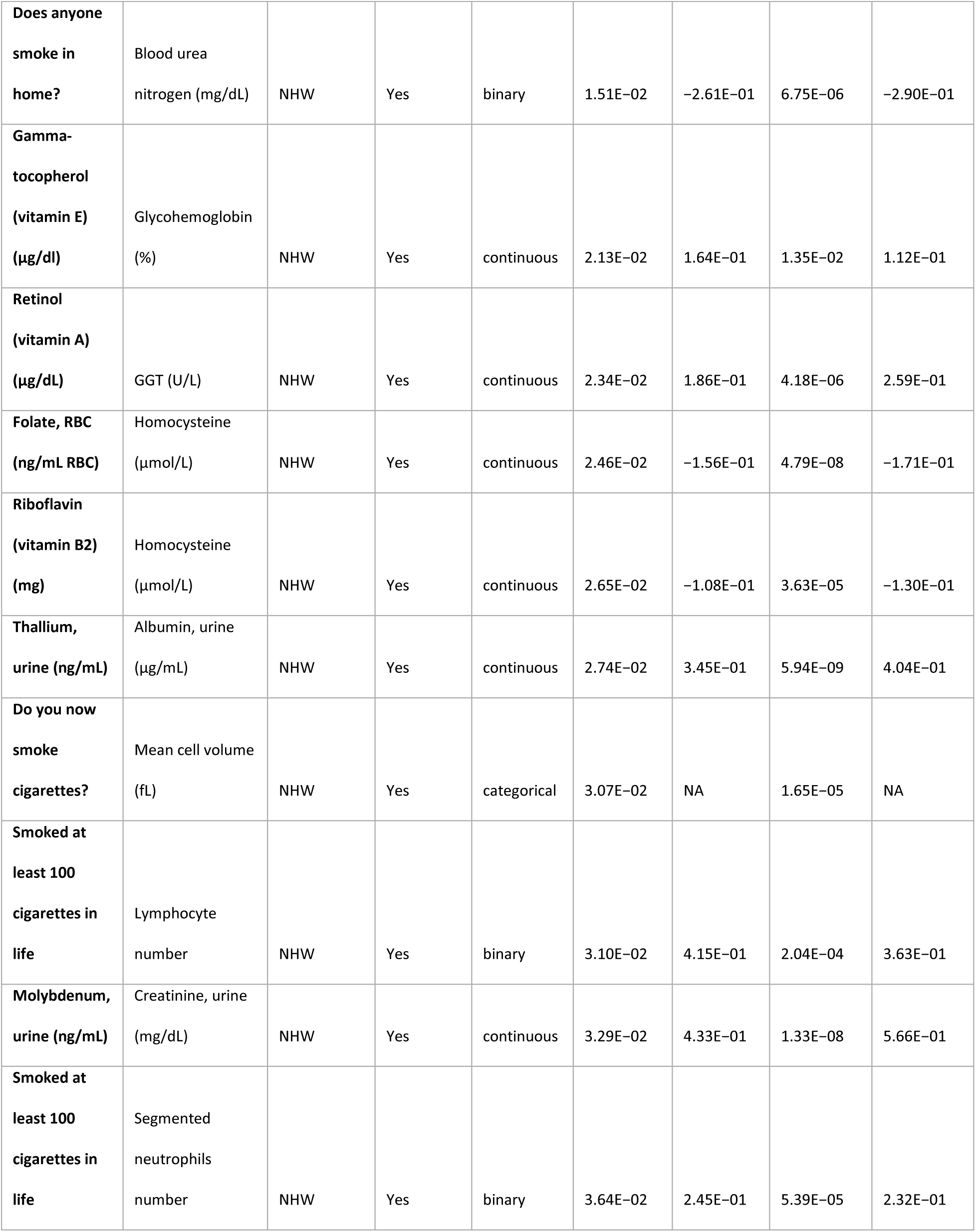
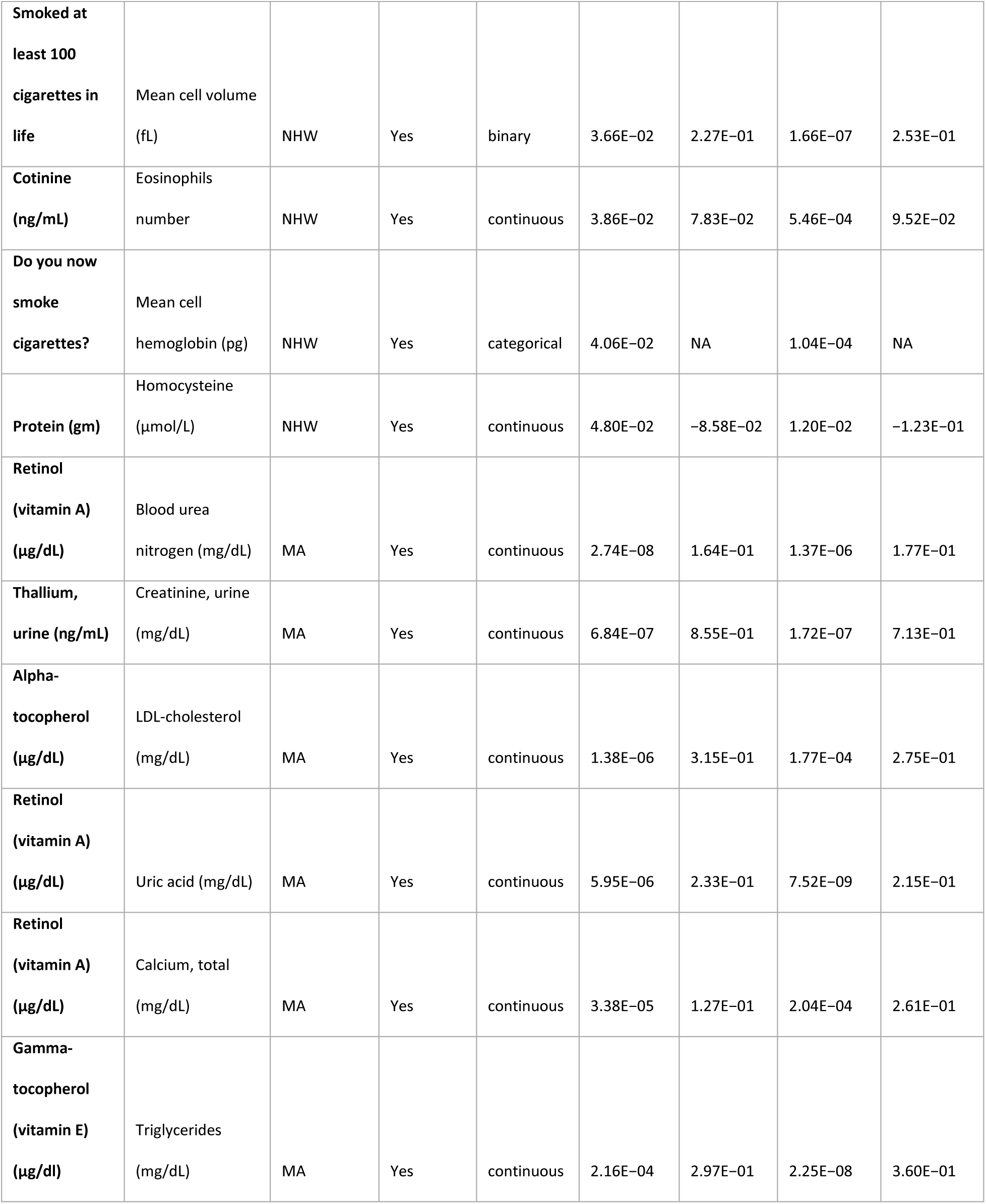
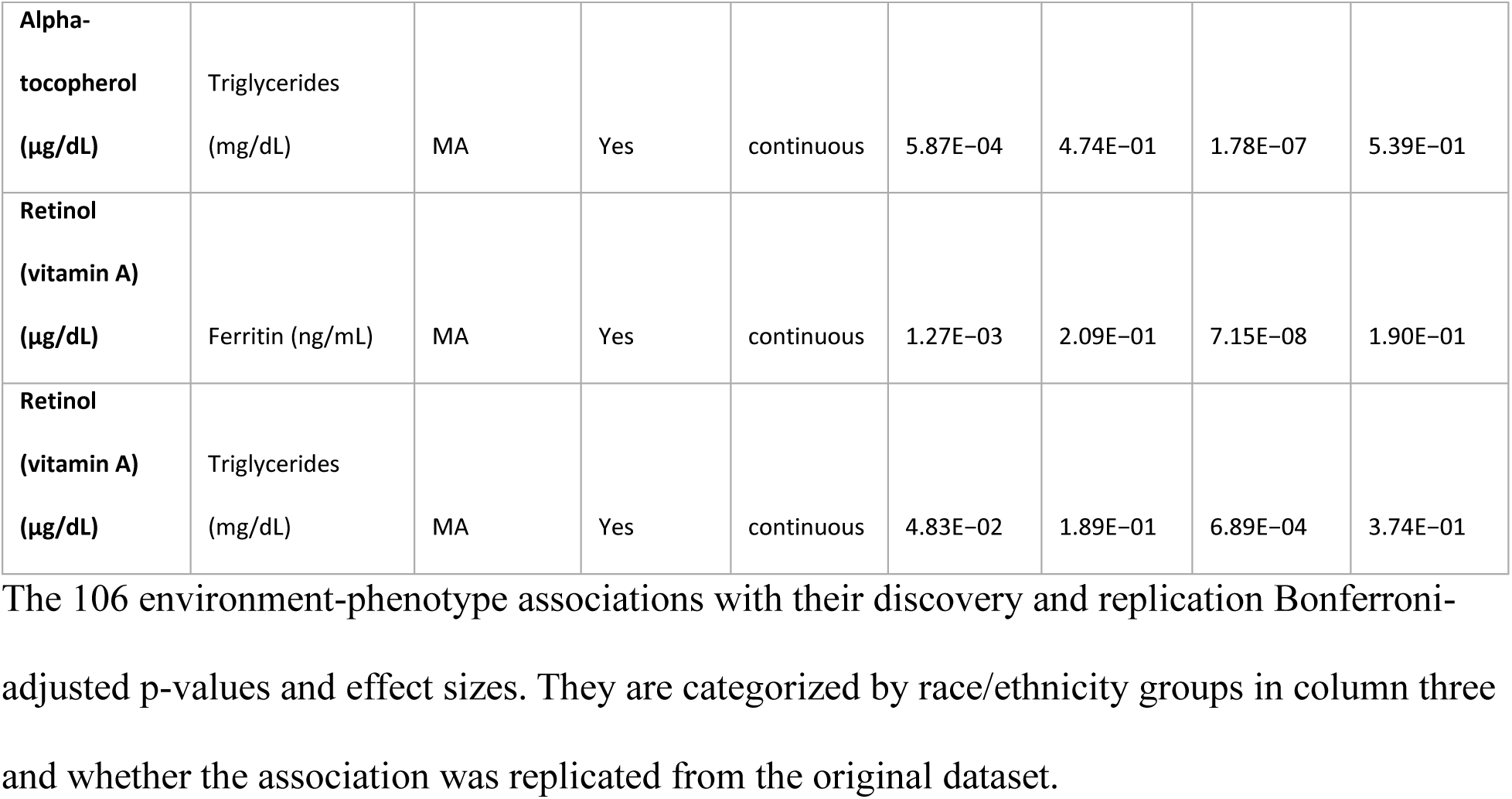
The stratified race/ethnicity PheEWAS results.

## DISCUSSION

PheEWAS assesses the effects of environmental exposures over a wide range of traits, which is important to gain an understanding of the interconnectedness of the environment and the phenome. In this study, we found 106 significant associations between exposures and phenotypes. Multiple exposures were significantly associated with numerous phenotypes, exemplifying the importance of exploring the impact of exposures at the phenome level.

Retinol (vitamin A) had the greatest number of cross-phenotype associations, with connections to 14 phenotypes (Table 1 & Fig. 2). Retinol is an essential, fat-soluble nutrient important in vision, growth, and metabolism^15–18^. It is obtained from plant and animal foods and by supplementation^15^. Although the mechanism is unknown, low levels of vitamin A impair iron metabolism^16^, while normal retinol levels have been associated with normal erythropoiesis and hemoglobin concentrations^17^. Thus, vitamin A deficiency is linked to anemia^16^. In this study, our results demonstrated retinol was associated with positive beta coefficients for various iron-related phenotypes, such as ferritin and total iron binding capacity (TIBC), as well as with hemoglobin concentration, reaffirming the role of retinol in iron metabolism and erythropoiesis. Higher retinol levels are also implicated in metabolic syndrome^18^. Because retinol is fat-soluble, it increases with higher body fat. Previous studies found that higher levels of vitamin A, uric acid, and gamma-glutamyl transferase (GGT) are associated with a higher risk of developing metabolic syndrome^18^. Therefore, the positive associations between retinol and uric acid, GGT, and triglyceride levels in our study suggest that retinol is part of a complex network that responds to elevated triglycerides through increased oxidative stress^18^. More specifically, GGT is a liver enzyme that serves as a biomarker for various hepatic and systemic conditions^19^. Elevated GGT levels have been associated with multiple exposures and substances, indicating its role in diverse metabolic and toxicological pathways where we observed associations with exposures like “drinks(s) per day”^20^ and cadmium^21^ while being associated with retinol. Although notably, GGT is not specific to lipids: it is similarly elevated after alcohol intake and with exposures to toxic metals including cadmium. The parallel associations we observe therefore reflect a united underlying oxidative-stress pathway that can be triggered by diverse metabolic and toxicological stressors.

Similarly, our results noted that alpha-tocopherol and gamma-tocopherol had cross-phenotype associations with positive beta coefficients related to lipid metabolism, including cholesterol and triglycerides. Vitamin E is a group of eight fat-soluble molecules, consisting of four tocopherols and four tocotrienols ^22^. The alpha- and gamma-tocopherols are ingested from food and supplements^23^. Alpha-tocopherol is usually obtained from olives and sunflower oils, and gamma-tocopherol is found in soybeans and corn oil^23^. Previous studies have found associations between alpha-tocopherol and gamma-tocopherol with triglycerides^24,25^ (Fig. 3). The fat-soluble tocopherols are transported in plasma by lipoproteins and erythrocytes; therefore, the associations between tocopherols and lipoproteins may reflect their joint transport^26^. We noticed that the phenotype serum homocysteine was associated with a positive beta coefficient for gamma-tocopherol but had a negative beta value for alpha-tocopherol. Homocysteine is reported to cause vascular damage by enhancement of lipoperoxidation, and alpha-tocopherol prevents the damage because of its fat-soluble antioxidant properties^27^. Although our PheEWAS found an association with a positive beta value for gamma-tocopherol and homocysteine, previous studies found inverse associations between alpha- and gamma-tocopherols and homocysteine^28^. Recent studies have also found that while alpha-tocopherol is associated with positive health outcomes, gamma-tocopherol may have a negative effect^28,29^. Although the mechanisms that underlie differences between tocopherol isoforms are unknown, these differences may result from their cell signaling functions. Multiple genes are differentially regulated by the tocopherol isoforms, including metabolic pathways genes^30^. Our results suggest different roles for alpha- versus gamma-tocopherol.

Cross-exposure associations with negative beta coefficients between homocysteine and the exposures riboflavin (vitamin B2), vitamin B6, and folate (vitamin B9) were also identified. The metabolism of homocysteine involves the demethylation of methionine with the aid of B-vitamins^31^. When homocysteine is broken down by the B-vitamins, it can convert into two substances. One substance is methionine, an essential amino acid for creating proteins, and the other is cysteine, a nonessential amino acid that reduces inflammation by communicating with immune cells^31^. B-vitamin supplements can increase the conversion of homocysteine into methionine and cysteine, thereby decreasing homocysteine in the blood. Dietary recommendations for folate and vitamin B12 take into account their association with homocysteine^32^. Adequate intake of B-vitamins, especially folate, is crucial for maintaining healthy homocysteine levels in the blood^32^. High homocysteine levels are associated with increased risks of cardiovascular diseases, including stroke and ischemic heart disease^30,31^. Research suggests that populations with lower-than-recommended B-vitamin intake, such as those in Sweden and Norway, often have elevated homocysteine levels^32^; thus, the associations of folate and other B-vitamins with homocysteine could result from a vitamin deficiency^31^. However, more research is required to determine whether there is a causal relationship between cardiovascular disease (CVD) and homocysteine and whether vitamin supplements to lower homocysteine in the blood affect CVD^33^.

Moreover, the associations between riboflavin (vitamin B2) and uric acid found in this PheEWAS extend this interconnected framework. Riboflavin plays a critical role in oxidative stress modulation, and its tandem relationship with uric acid—both an antioxidant and pro-oxidant—further emphasizes shared pathways in oxidative metabolism^34,35^. These associations are anchored in the well-established relationship between riboflavin and homocysteine, where riboflavin acts as a cofactor in homocysteine metabolism, promoting its breakdown into methionine and cysteine^34^. When homocysteine builds up, it creates harmful oxidative stress, and uric acid helps neutralize those damaging molecules. Vitamin B2 (riboflavin) is needed to make the enzymes that both break down homocysteine and produce uric acid, linking the two processes. Similarly, folate (vitamin B9), which is crucial for DNA synthesis and erythropoiesis, is also closely linked to homocysteine metabolism, underscoring its role in maintaining cardiovascular and hematological health^36,37^.

Other inverse associations with homocysteine were found in this study for several micronutrients, including potassium, which controls nerve function and muscle contraction^38^. Atrial myocytes, which form the muscular walls of the heart, use potassium to regulate their contractile function; decreased potassium can lead to atrial fibrillation^39^. Homocysteine at 50 µmol/L and 500 µmol/L results in a significant decrease in potassium movement in human atrial myocytes^39^. The mechanism by which homocysteine directly affects myocytes is unknown; however, our finding that homocysteine was inversely associated with potassium supports its possible effect on myocytes^39^. Our results are consistent with prior studies showing associations between elevated homocysteine levels and adverse health outcomes, particularly in the context of cardiovascular disease and cardiac arrhythmias, though these specific outcomes were not directly evaluated in our phenotype set. Overall, our results indicate a trend of adverse outcomes associated with increased homocysteine.

Cotinine, a biomarker for tobacco smoking exposure^40^, had 12 cross-phenotype associations. Smoking behavior is a risk factor for heart disease, lung cancer, stroke, and chronic obstructive pulmonary disease (COPD)^41^. Increased cotinine levels and smoking behaviors are associated with inflammation and an increase in WBCs^42,43^. We found associations with a positive beta coefficient between cotinine and smoking-related behaviors with several WBC counts. This supports the hypothesis that smoking behavior can lead to systemic inflammation that increases WBC^44^. This is also supported by our finding of an association with positive beta value between cotinine levels and C-reactive protein, an inflammatory biomarker^45^. In our study, cotinine was also associated with increased RBCs. It has been hypothesized that, due to carbon dioxide being carried by cigarette smoke, increased hematocrit and hemoglobin levels compensate to maintain oxygen transport^45^. Our analysis detected positive directions-of-effect for cotinine levels and other smoking-related behaviors with hemoglobin and hematocrit, indicating an increase in RBCs with smoking exposure^46^.

Our analysis showed that serum cadmium levels had 11 cross-phenotype associations, including WBC and RBC phenotypes. Cadmium is a toxic heavy metal that accumulates in plants, particularly tobacco and edible crops grown in cadmium-contaminated soil and water. Therefore, exposure to cadmium is often via smoking and/or food^47^. Exposure to cadmium causes oxidative stress and affects the kidneys, liver, bone, and lungs^12,48^. We observed that cadmium and cotinine shared similar associations, which may result from cadmium exposure via tobacco smoke^13,49,50^. There was a positive direction of effect between cadmium and mean RBC volume. However, WBC counts also increased with cadmium. *Ciarrocca et al.,* found that, in workers exposed to cadmium, there was a positive correlation between cadmium and lymphocyte count. Cadmium induces apoptosis of lymphocytes, thereby modulating the immune system^52^. Our results show similar associations between cotinine and cadmium, which could reflect their co-occurrence in tobacco. However, because cadmium is also an industrial pollutant, it could affect WBC and RBC counts independent of cotinine^53^. As we learn more about the effects of cadmium exposure on multiple phenotypes, we will be able to inform treatment or prevention of diseases related to excess cadmium.

When we analyzed each race/ethnicity stratum, we identified many significant associations that were shared with the unstratified analysis. Alpha-tocopherol with triglycerides, retinol with triglycerides, retinol with uric acid, retinol with blood urea nitrogen, and gamma-tocopherol with triglycerides were the top five significant associations for exposures and phenotypes, respectively, in the unstratified and stratified analyses. Race/ethnicity is often adjusted in linear regression models as a covariate which precludes investigation of population-specific associations^54^. To determine if any associations varied by population, we assessed differences between the stratified and unstratified results. Eleven associations were found in the stratified analysis that were not significant in the unstratified dataset, nine of which were identified in the NHW sample. The NHW sample was larger than the NHB and MA samples, which may have driven the higher number of observed associations within that stratum.

Two associations were only observed in the MA stratum, an association with a positive direction-of-effect involving cadmium exposure and the creatinine phenotype, and an association with an inverse direction-of-effect between platinum exposure and eosinophil count. The positive direction-of-effect of cadmium and creatinine, implies the potentially toxic effects of cadmium exposure on health. A previous study found that Mexican Americans living in urban and agricultural areas had higher urinary and blood cadmium concentrations compared to other populations^55^. This elevated exposure was linked to increased risk of kidney damage, as reflected in higher serum creatinine levels^55^. These findings demonstrate the need for further study of cadmium exposure in this population.

Only the NHW stratum showed a positive direction-of-effect between urinary lead and urinary creatinine. Lead toxicity is determined by urine lead concentration per gram of creatinine^56^. Lead toxicity produces reactive oxygen species (ROS) causing oxidative stress^56,60^. Lead-induced oxidative stress involves the generation and removal of ROS in cells and tissues, which damages membranes, DNA, and proteins^56^. An increased intake of antioxidants can decrease susceptibility to oxidation in those tissues^27^. Supplementation with beta-carotene decreases homocysteine in the blood and has been shown to reduce oxidative stress in chronically lead-exposed workers^57^. Beta-carotene is a carotenoid that protects the body from free radicals; when homocysteine is broken down, beta-carotene increases antioxidant production, reducing oxidative stress^27,57^. The inverse relationship we observed between trans-beta-carotene and homocysteine in the NHW sample supports these findings.

Using PheEWAS, we leveraged data from the NHANES to interrogate the complex relationship between the exposome and phenome. We replicated several associations previously described in the literature and identified novel relationships as well. Limitations to this study should be noted. 1) Like genome-wide association studies, our study provides limited information on the mechanisms linking the exposures and phenotypes; however, these associations can provide novel hypotheses for future studies. 2) The nature of the NHANES provides only cross-sectional snapshots of the exposure and phenotypic associations during adult stages, thereby missing dynamic longitudinal changes in the exposome and their phenotypic effects. In the future, we hope to incorporate longitudinal data and methods in phenomic environment-wide analyses. 3) Because classifying a characteristic or an analyte as a phenotype or exposure can be an arbitrary decision, we were strict in our classifications. We combed the NHANES dataset and dictionary to accurately identify a) phenotypes based on blood and biochemistry lab measurement health outcomes and b) exposures based on their definitions as exogenous factors. Furthermore, the direction of causality was not explored in this study; in some cases, our defined exposures could be affected by an individual’s health status, and in other cases, both exposure and phenotypes could be affected by factors that were not assessed by NHANES. This illustrates the challenges of the high-throughput analysis of exposome and phenome data. 4) Our interpretation of the PheEWAS requires further downstream hypothesis testing. A possible follow-up study could use a more restrictive statistical model that incorporates covariates unique to one of the significant exposure-phenotype associations that we would not necessarily control for in a generic EWAS model. Re-sampling the data using methods like bootstrapping will also improve the interpretation of results.

PheEWAS is a novel approach that connects the exposome with the phenome. As characterization of the exposome becomes more possible with a diverse array of technologies and assessment measures, its complex relationship with the phenome will be further revealed. When applied to a wider range of these exposures, and across multiple timepoints, PheEWAS will enable increased knowledge of the impact of the environment on human health and the ability to identify those most vulnerable to developing adverse health outcomes.

## METHODS

### National Health and Nutrition Examination Survey (NHANES)

This PheEWAS identified associations between multiple phenotypes and environmental exposures in adults using data from NHANES, which evaluates the health and nutritional status of adults and children in the United States^14^. The survey consists of dietary, socioeconomic, and general health data obtained through questionnaires and clinical laboratory measurements. These are data collected annually by the Centers for Disease Control and Prevention (CDC) public health policies^14^.

We split the NHANES data into discovery and replication cohorts to demonstrate replicability within the dataset. We excluded individuals under age 18, leaving 18,850 individuals, 970 exposure variables, and 56 phenotypes across four survey periods (1999−2000, 2001−2002, 2003−2004, 2005−2006) prior to quality control (QC). The participants in the 1999−2000 and 2001−2002 datasets comprised the discovery dataset (N = 9,063; 52% female, 47% NHW, 19% NHB, 25% MA, 5% other Hispanic, and 3% other ethnicities). The participants in the 2003−2004 and 2005−2006 datasets comprised the replication dataset (N = 9,874; 52% female, 49% NHW, 23% NHB, 21% MA, 3% other Hispanic, 1% other ethnicities) (Table 3). Demographic information was obtained through participant interviews or physical examinations at the mobile exam center^58^.

**Table 3:**
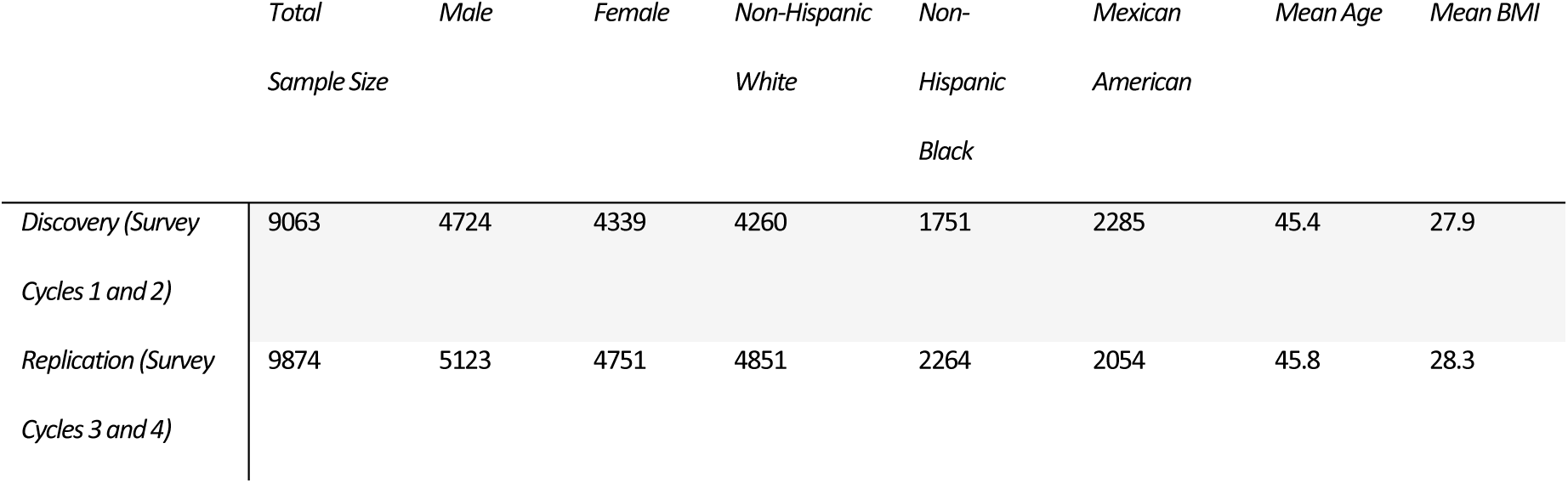
Demographics of the discovery and replication samples.

### Selection of phenotypes and exposures

The phenotypes were selected from the blood and biochemistry laboratory measurements recorded in the NHANES Dataset Explorer^59^. Exposures were obtained from questionnaires, examinations, and other laboratory measurements, and comprised the following categories: alcohol use, bacterial infection, cotinine, dialkyl, dioxins, food component recall, furans, heavy metals, hydrocarbons, immunizations, nutrients, occupations, polychlorinated biphenyls, pesticides, pharmaceuticals, phthalates, phytoestrogens, sexual behavior, smoking behavior, smoking family, social support, street drugs, supplements, and volatile chemicals. To better summarize the above exposure categories when listing cross-phenotype results, we aggregated them into the following: dietary supplements (nonmetals and metals), metal pollutants, vitamins, alcohol, smoking, and drugs and chemicals. We utilized phenotype in the biochemistry or blood sample category in the NHANES data dictionary. For blood or biochemistry measurements, the phenotypes were divided into categories for assessing multi-functional effects: WBCs, iron measurements, amino acids, RBC measurements, lipids, metabolic products, enzymes, and blood proteins. All blood or biochemistry markers were deemed a measurable characteristic, as they reflected the physiological state of an individual that could be influenced by environmental factors. For a full list of phenotypes and exposures, see Supplemental Table 3.

### Quality control of phenotypes and exposures

Before QC, 970 environmental variables and 56 phenotypes from the NHANES dataset were considered for the discovery dataset. The QC pipelines, protocols, and covariate adjustments for the discovery and replication datasets were identical.

#### Data filtering and sample/variable-based exclusion

Using CLARITE Software^11^, environmental exposures were classified as binary, categorical, or continuous. All phenotypes were continuous. Covariates included for adjustment were age, sex, socioeconomic status (SES), BMI, self-reported race/ethnicity, and data release cycle year. Participants missing a covariate value were excluded, leaving 9,063 samples in the discovery and 9,874 in the replication datasets. Categorical and binary variables that had less than 200 samples in any category were dropped^60^. To determine whether 200 was a sufficient sample size, we simulated over 1,000 observations and found the optimal power of 80% with detection of effect sizes greater than 0.4 (Supplemental Fig. 1 a, b).

#### Data distribution and transformation of phenotypes

Skewness was calculated and histograms for all phenotypes were visualized for all phenotypes using CLARITE Software (Supplemental Fig. 2), and a log transformation was performed on skewed phenotypes. Phenotypes that had a normal distribution and a skewness value of −0.5 to 0.5 were not transformed. Thirteen discovery phenotypes were normally distributed and were not transformed, whereas 45 phenotypes were log-transformed (Supplemental Fig. 2 a-c). For negatively skewed phenotype *p*, values of *p* were adjusted by subtracting them from the constant [1 + max(*p*)]. The new values were log-transformed, and once again adjusted by subtracting them from the new [1 + max(*p*)] to return to their original orientation^61^. Continuous phenotypes for which more than 90% of samples had a value of zero were removed. After QC, 326 exposures (277 continuous, 45 binary, and four categorical variables) and 55 phenotypes remained for analysis. Further, to maintain sufficient sample size, we removed any exposure-phenotype combination with a total sample size or category size (if categorical or binary) < 200, leaving 17,909 out of the 17,930 exposure-phenotype pairs with a total sample size and, if applicable, category size > 200 that was included in the PheEWAS.

#### Lipid phenotypes

When a participant reported taking a statin (atorvastatin, simvastatin, pravastatin, or fluvastatin), the low-density lipoprotein (LDL)-cholesterol value was divided by 0.7 to estimate the free LDL cholesterol level prior to statin treatment, as recommended by the Global Lipids Genetics Consortium^62,63^.

### Survey weights

We included survey weights in our analysis to account for the complex survey design of NHANES as recommended^64^. NHANES selected participants from primary sampling units (PSU) and strata while oversampling typically underrepresented groups, such as minorities and the elderly. Survey cycles one and two (1999–2002) were assigned four-year weights as defined by the population estimates used in the 1990 and 2000 census years^64^. In contrast, survey cycles three and four (2003–2006) had two-year weights from which the four-year weights were calculated by dividing their respective two-year weight in half^65^.

### Phenomic environment-wide association study

Fifty-five phenotypes and 326 exposures that passed QC were included in the PheEWAS (Supplemental Table 1). All PheEWAS models were tested using CLARITE Software. We used a general linear model that was adjusted for sex, self-reported race/ethnicity (Non-Hispanic White, Non-Hispanic Black, and Mexican American), body mass index (BMI), SES, age, and survey year. Complex survey design data was passed into the model including the strata, cluster, and weights for specific variables. A Z-score transformation of continuous or binary exposures and continuous phenotypes was used to standardize the beta coefficients and allowed for a direct comparison of effect sizes across different exposure-phenotype associations^10^. If the predictor was continuous or binary, CLARITE reports the beta coefficient and p-value from the general linear model. For categorical predictors, CLARITE compares the model with and without the categorical predictor using a likelihood ratio test (LRT) to assess whether the inclusion of any (or all the) categories of the predictor improves the model fit. Significant exposure-phenotype associations with a false discovery rate (FDR) p-value < 0.10 in the discovery were considered for replication. Replication criteria for exposure-phenotype associations were 1) an FDR p-value < 0.10 in the replication dataset and 2) the same direction of effect between subsets.

Three race/ethnicity strata provided by the NHANES questionaries—Non-Hispanic Black, Mexican American, and Non-Hispanic White—had sufficient sample sizes to include in the stratified analysis. Here, we applied the same discovery and replication pipeline as for the unstratified PheEWAS. In the stratified PheEWAS, we accounted for the complex survey design for NHANES similar to the unstratified PheEWAS. The same strata, PSUs, and sampling weights were used. However, stratifying the NHANES data introduced strata with only a single PSU, which can occur in subsets of data^66^. In CLARITE Software, the traditional variance calculation must be adjusted; therefore, we used the ‘adjust’ option, which was incorporated from the survey package in R. This option provides a conservative estimation of variance by using population residuals rather than stratum residuals, ensuring a more conservative estimate of variance in complex survey designs^66^.

## Data Availability

More information on the data is available on Dryad (https://doi.org/10.5061/dryad.d5h62)^59^

All code, QC files, and result files can be found on GitHub (https://github.com/HallLab/PheEWAS_paper/tree/main)

## Supporting information

Supplemental Figures And Table Descriptions

## Data Availability

Data Availability
More information on the data is available on Dryad (https://doi.org/10.5061/dryad.d5h62)
All code, QC files, and result files can be found on GitHub (https://github.com/HallLab/PheEWAS_paper/tree/main)

## Acknowledgements

This work is supported by the USDA National Institute of Food and Agriculture and Hatch Appropriations under Project #PEN04275 and Accession #1018544

## Author contributions

N.E.P., T.G.Z., and K.P. contributed the formal analysis, investigation, methodology, writing and editing the manuscript. J.Z. contributed the formal analysis, methodology, and editing the manuscript. D.C.C. contributed investigation, methodology, and editing the manuscript. M.A.H., contributed the conceptualization, formal analysis, investigation, methodology, writing and editing the manuscript, project administration, and supervision.

## Competing interests

The authors declare no competing interests.

## Notes

### Competing Interest Statement

The authors have declared no competing interest.

### Author Declarations

The National Health and Nutrition Examination Survey (NHANES) is a population survey implemented by the Centers for Disease Control and Prevention (CDC) to monitor the health of the United States whose data is publicly available in hundreds of files. This Data Descriptor describes a single unified and universally accessible data file, merging across 255 separate files and stitching data across 4 surveys, encompassing 41,474 individuals and 1,191 variables. The variables consist of phenotype and environmental exposure information on each individual, specifically (1) demographic information, physical exam results (e.g., height, body mass index), laboratory results (e.g., cholesterol, glucose, and environmental exposures), and (4) questionnaire items. Second, the data descriptor describes a dictionary to enable analysts find variables by category and human-readable description. The datasets are available on DataDryad and a hands-on analytics tutorial is available on GitHub. Through a new big data platform, BD2K Patient Centered Information Commons (http://pic-sure.org), we provide a new way to browse the dataset via a web browser (https://nhanes.hms.harvard.edu) and provide application programming interface for programmatic access. https://datadryad.org/dataset/doi:10.5061/dryad.d5h62

### Summary of Updates

This version of the manuscript has been revised to update figure legibility on page 34.

