## Supplemental Figures And Table Descriptions for "Phenomic environment-wide association study (PheEWAS) models complexity in the exposome"

**SUPPLEMENTARY FIGURES**

**Supplementary Table S1 (in text file). Overview of replicating results with Bonferroni-adjusted p-values.** 106 phenotype-environment associations are shown with their Discovery and Replication raw p-values and sample size.

**Supplementary Table S2 (in text file). Overview of replicating results with Bonferroni-adjusted p-values found in multiple racial/ethnic strata.** 106 phenotype-environment associations are shown with their Discovery and Replication p-values and sample size. The group column specifies which race-ethnicity strata the test originated: Non-Hispanic Black (NHB), Mexican American (MA), and Non-Hispanic White (NHW).

**Supplementary Table S3 (in text file).** Column 1 lists the 326 exposures used as the predictor variable for each individual test and column 3 are the 56 phenotype outcomes with corresponding CDC codes.

# a) b)

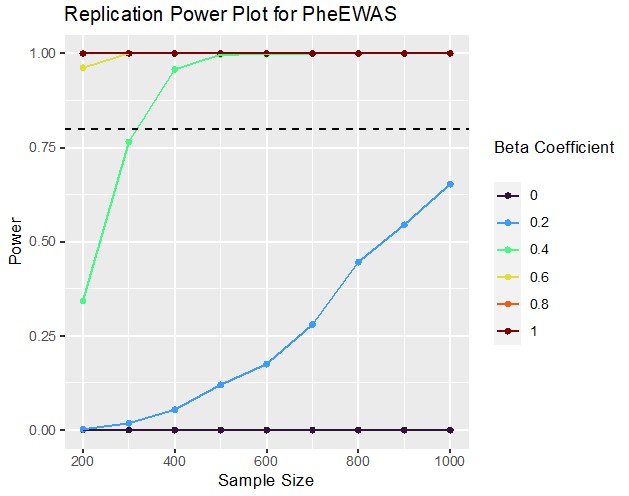

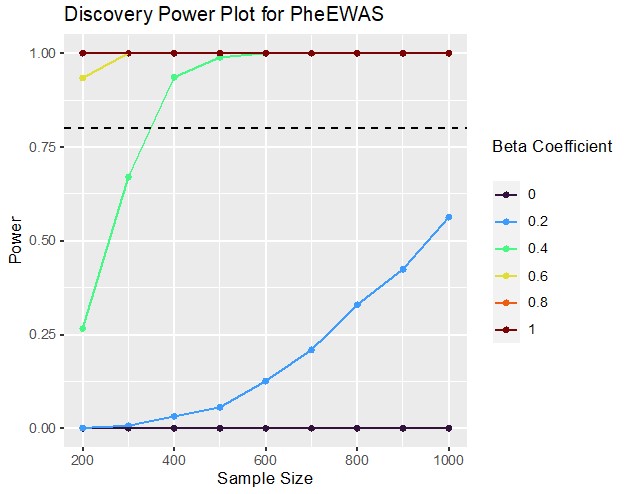

**Supplementary Figure 1.** Based off the linear regression model used in this study, two power plots a) Discovery and b) Replication display the sample size on the x-axis and the magnitude of power based on the beta coefficient on the y-axis. The key identifies the magnitude of beta coefficient, and the dotted black line signifies an 80% power threshold.

# A)

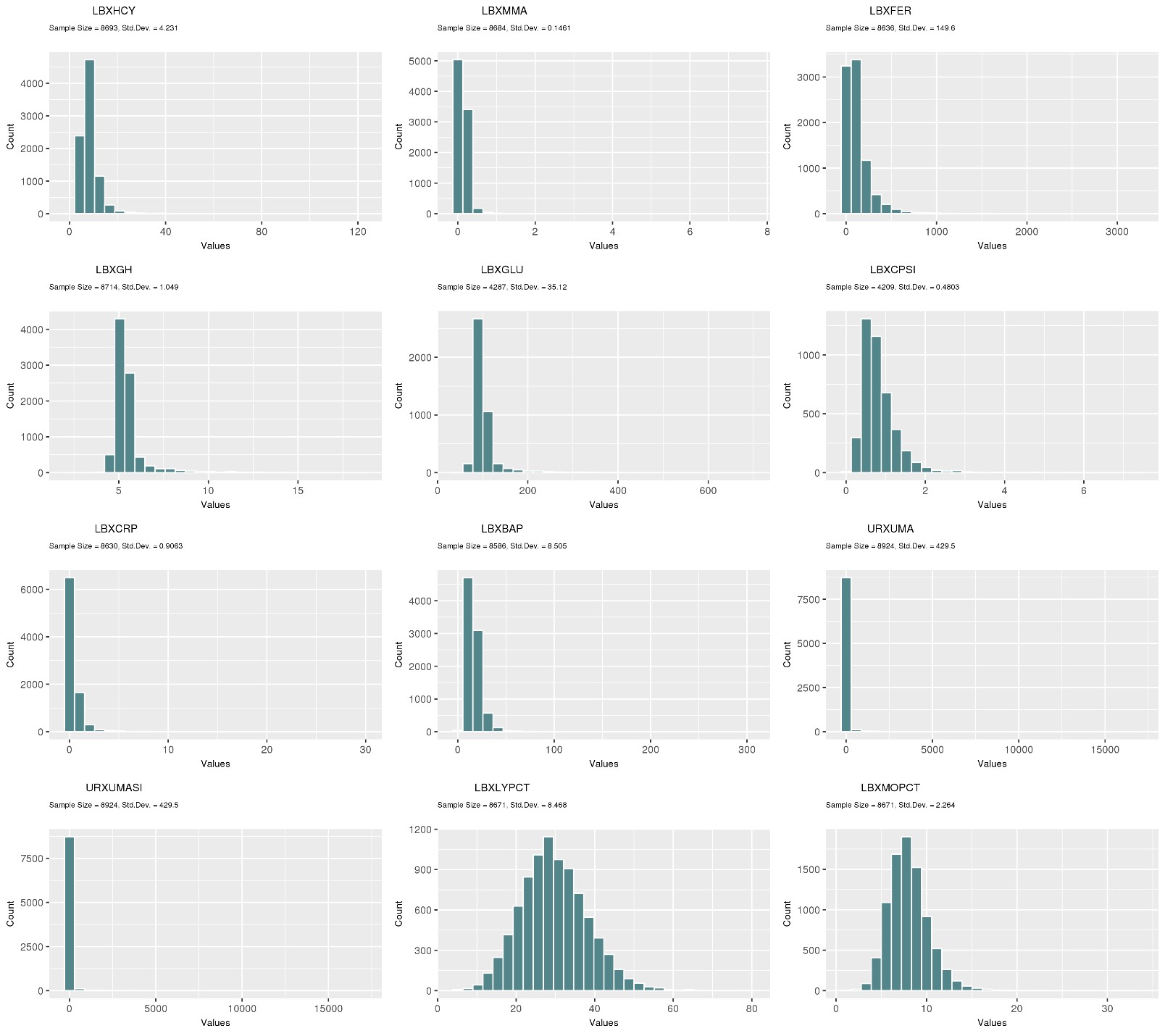

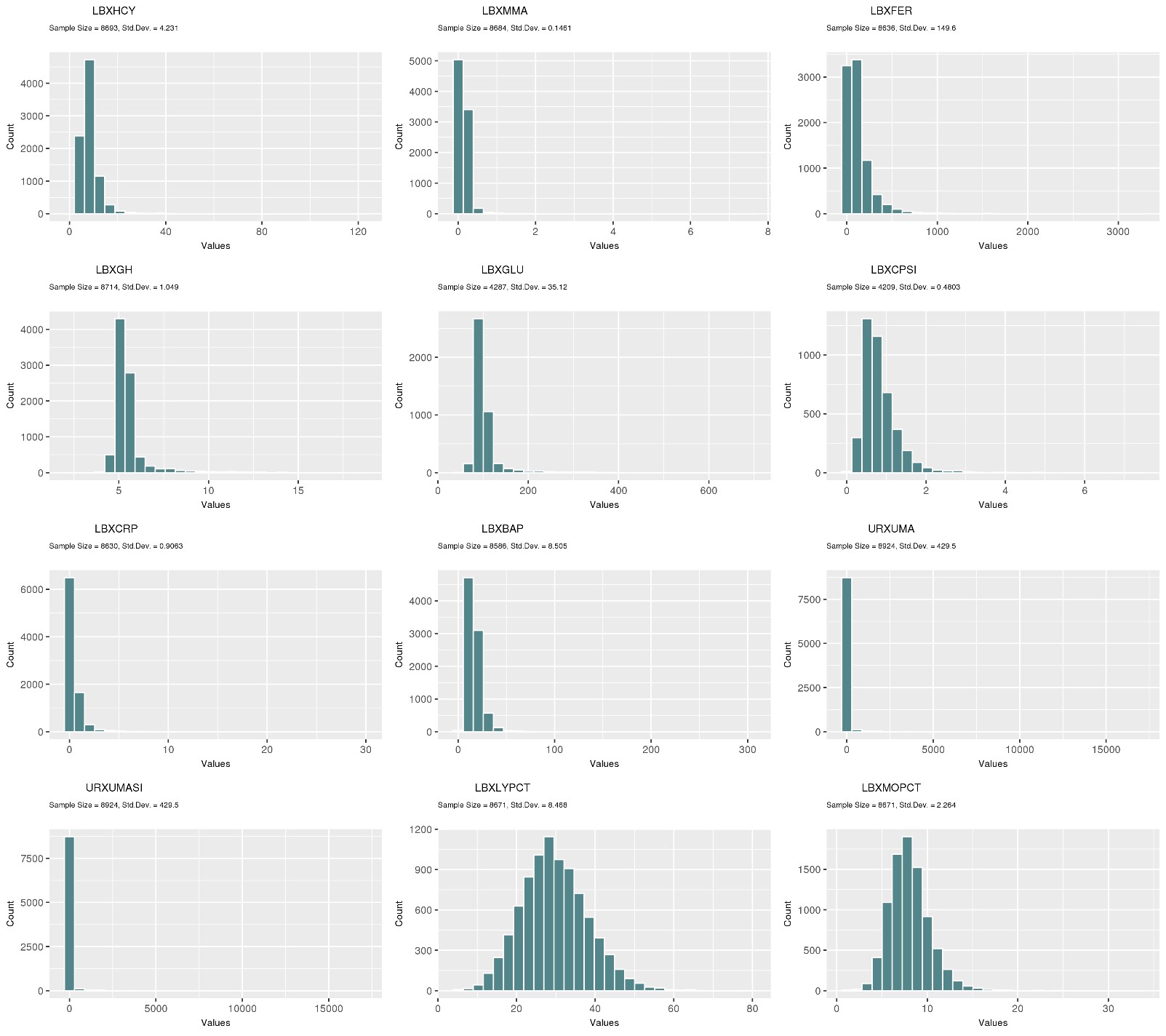

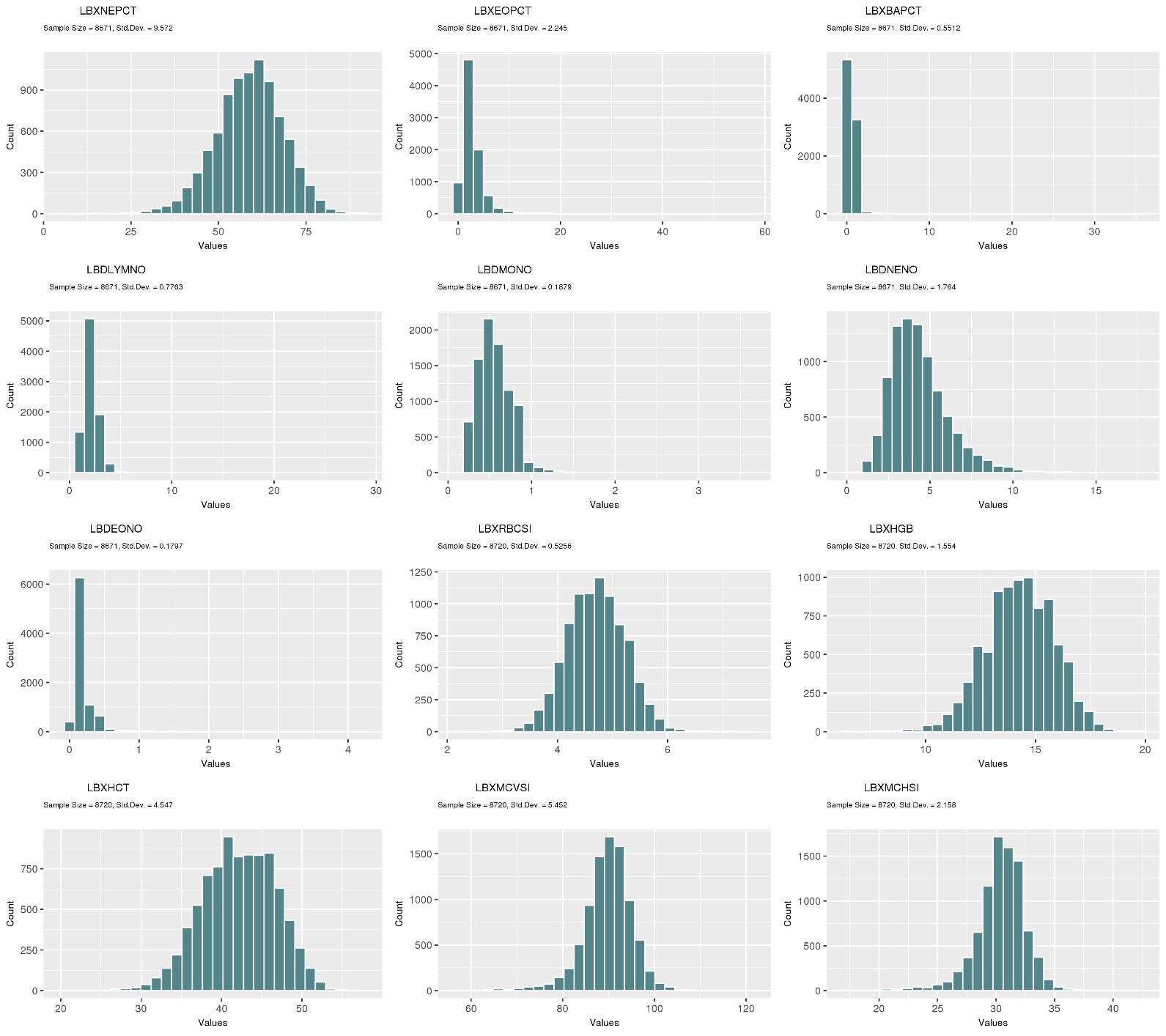

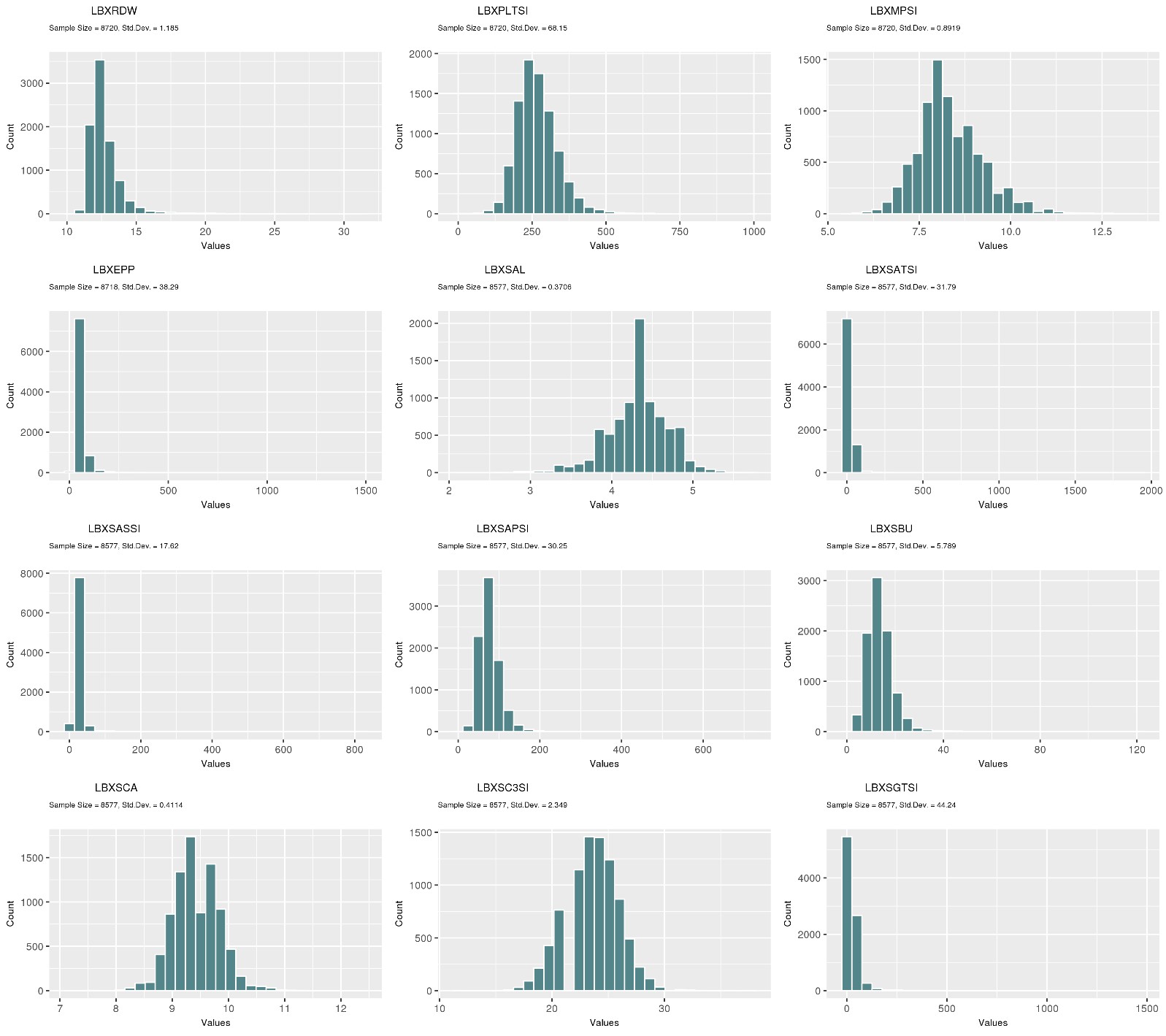

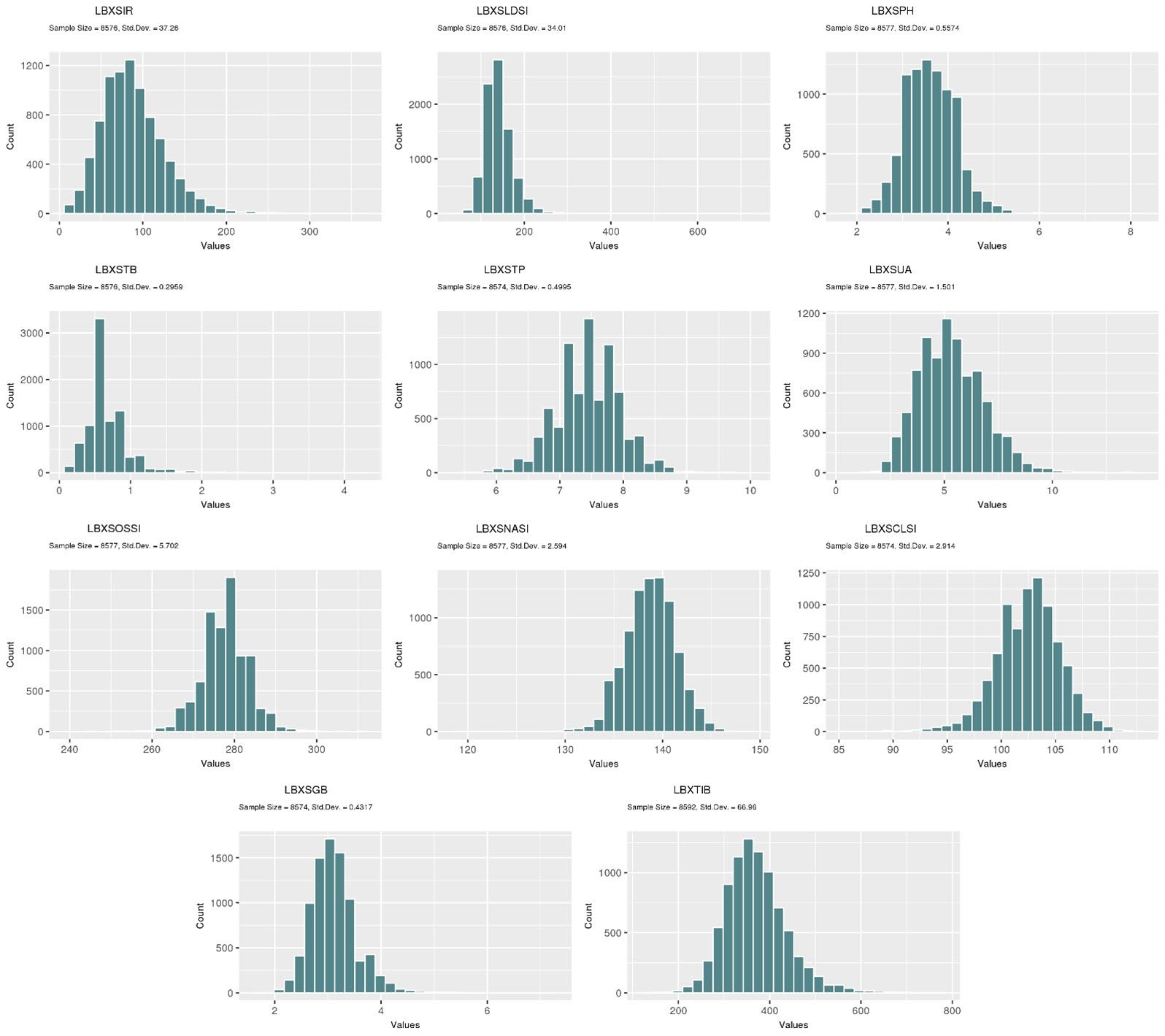

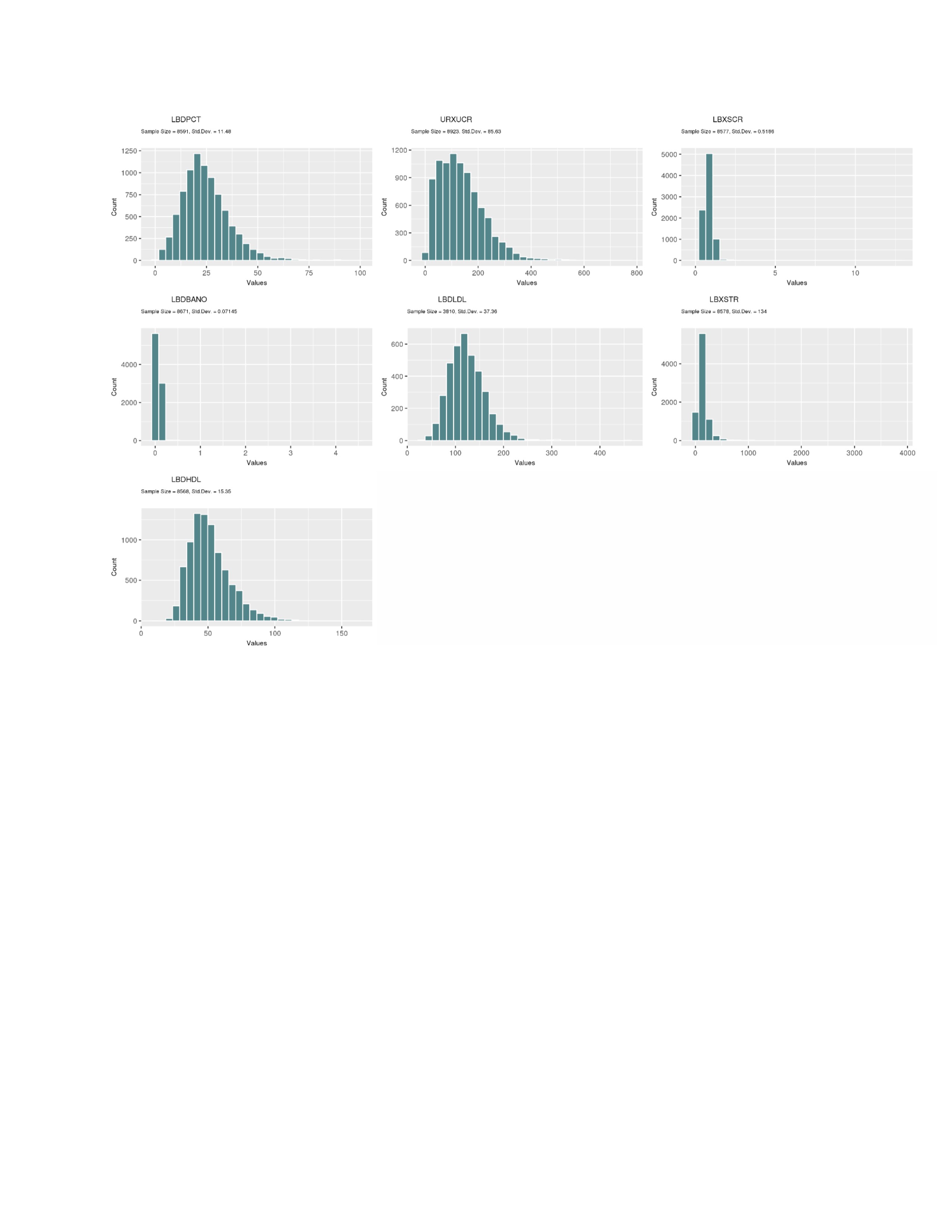

# B)

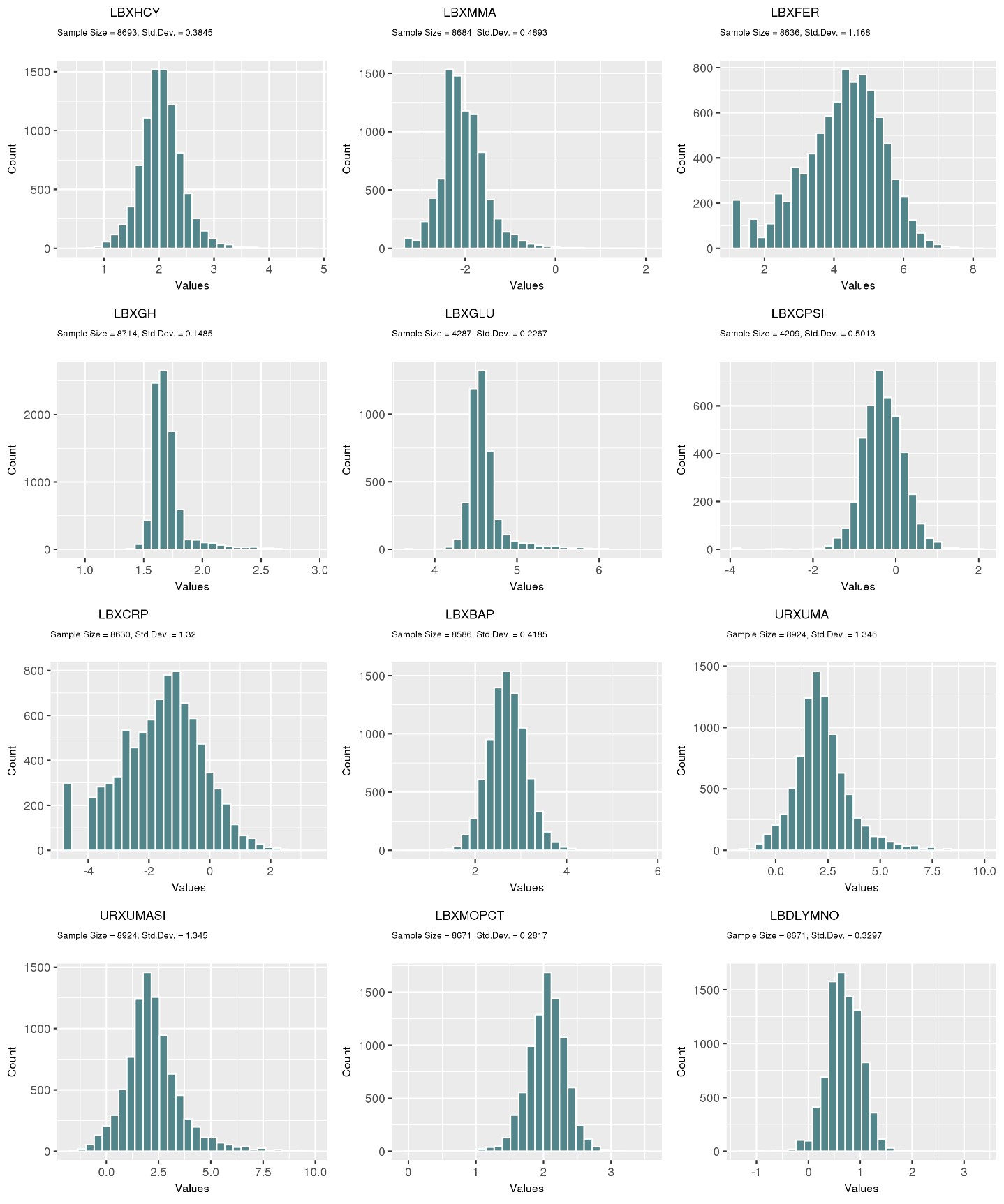

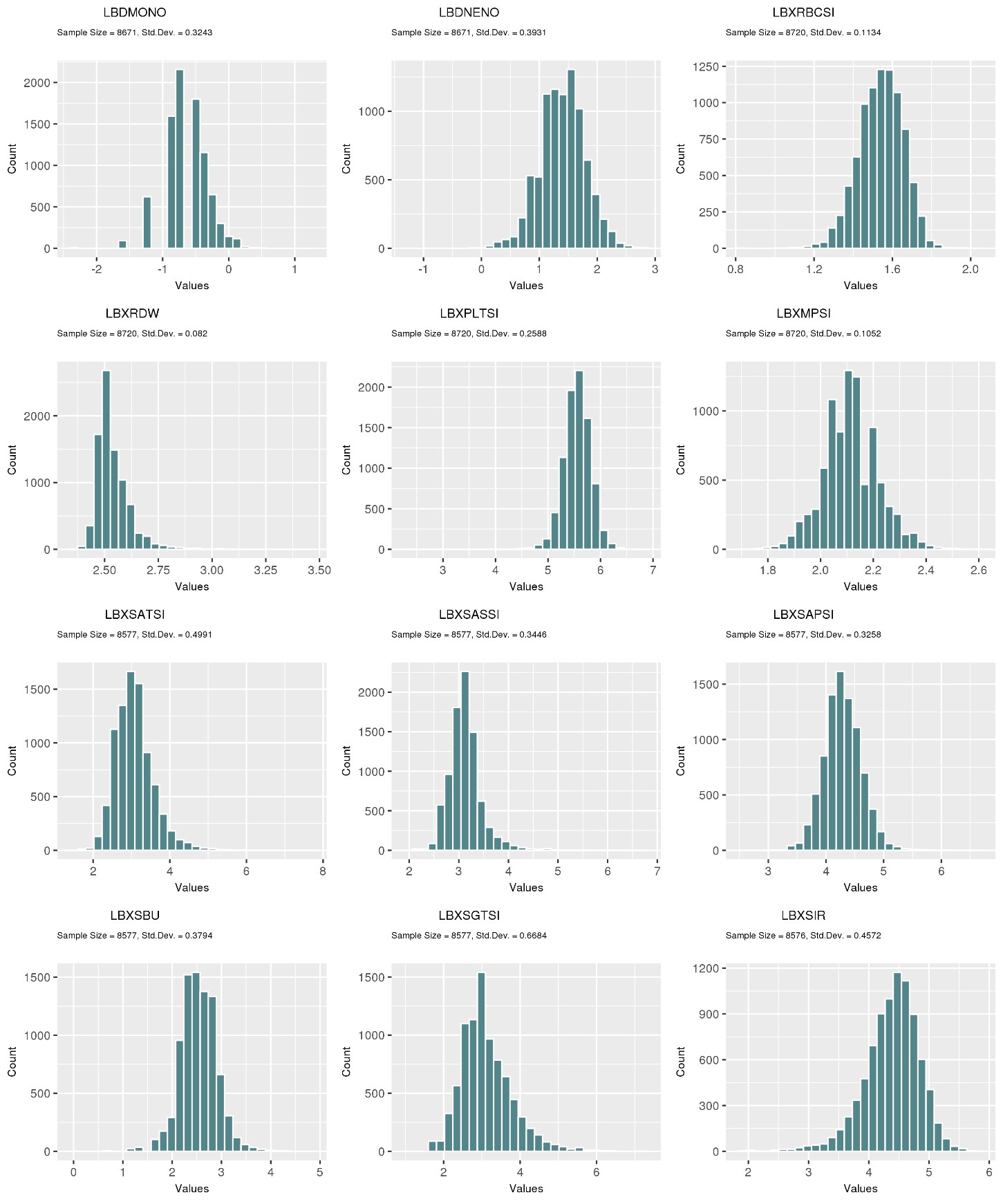

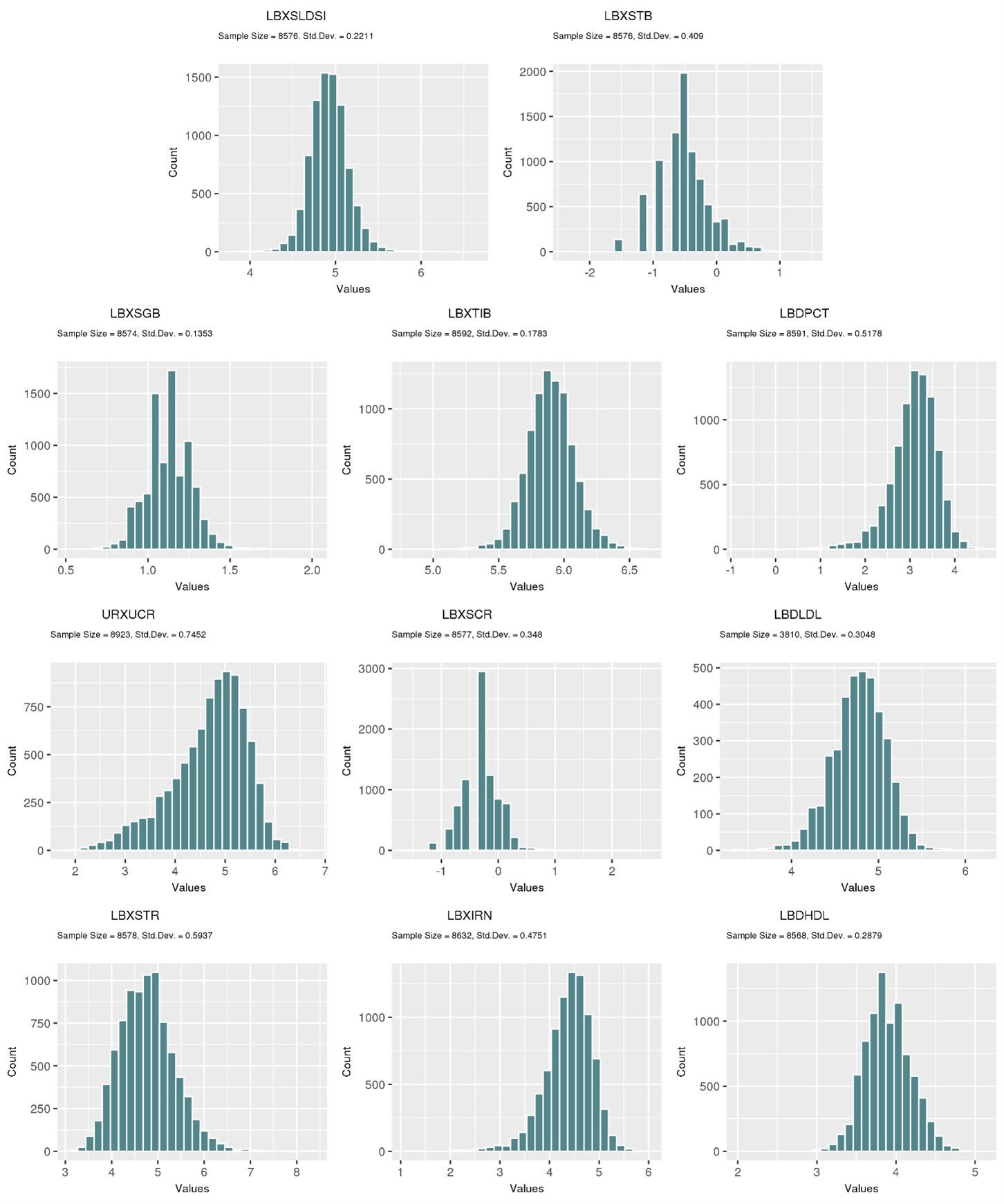

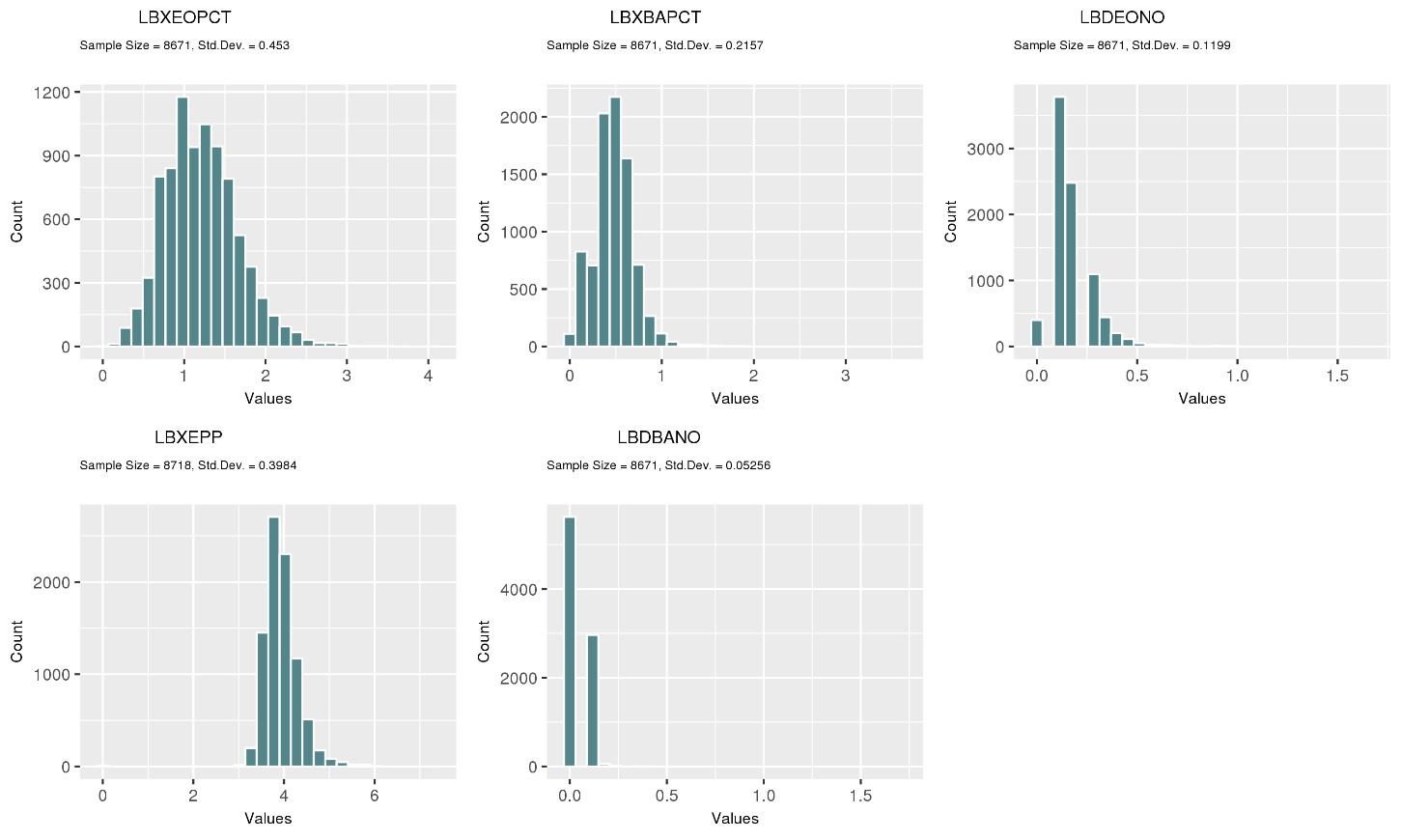

# C)

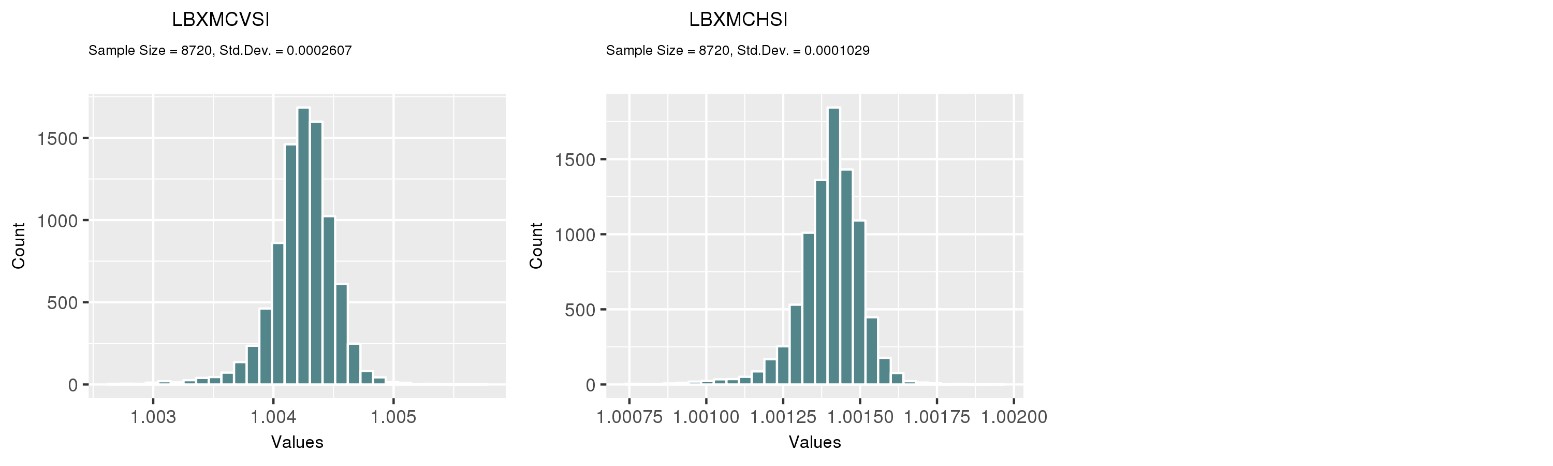

**Supplementary Figure S2.** A) Histograms displaying the distributions of the phenotypes from the Discovery dataset prior to transforming. Histograms after the B) positive and C) negative transformations of the phenotype values was implemented based on skewness in order to normalize the data.
